# LitBench: A Benchmark for Retrieval-Grounded Multi-Paper Evidence Synthesis in Epilepsy

**DOI:** 10.64898/2026.09.17.26363288

**Authors:** Alon Gorenshtein, Tianyu Zhang, M. Brandon Westover, Daniel M. Goldenholz

## Abstract

**Background:** Clinicians increasingly use AI systems to search **the** medical literature, but current benchmarks do not jointly **test** whether a response identifies the originating paper and its supporting passage. LitBench evaluates four dimensions: stated versus interpretation-requiring facts, the number and composition **of** competing papers, one-versus two-paper evidence, and refusal when evidence is absent.

**Methods:** LitBench comprises 1,980 open-access papers (1,000 epilepsy papers holding the answers, 980 decoys from unrelated fields) and 9,472 human-reviewed facts, giving 2,188 single-paper questions and 170 requiring a fact from each of two papers. Difficulty rose by burying the answer among up to 2,000 distractors, then again over live PubMed Central. The same questions were then asked with the answering paper removed, so that refusal was the correct response. Four systems were tested: Gemma-4B, Gemma-12B, Sonnet-5, and DeepSeek-V4-Flash (refusal conditions only). Three model judges scored each answer by majority; two-paper questions counted only when both facts were found.

**Results:** Across fixed-corpus single-paper conditions, accuracy ranged from 55.9% to 71.9% for Gemma-4B, 61.2% to 75.5% for Gemma-12B, and 90.0% to 91.3% for Sonnet-5. Similar epilepsy papers were harder than mixed candidate sets for both Gemma configurations. Two-paper accuracy was 5.7%, 17.1%, and 37.8%, respectively. With evidence absent, Gemma-12B refusal fell from 88.3% to 39.9% as candidate sets grew; Gemma-4B almost never refused. Sonnet-5 refused on 94.9% of sampled single-paper and all sampled two-paper cells. DeepSeek-V4-Flash also refused frequently, but on 80.3% of matched answer-present controls.

**Conclusions:** No system was reliable across every axis tested: accuracy fell as competing papers were added, interpretation-requiring facts were harder than stated ones, two-paper synthesis was rarely achieved, and only some systems recognized absent evidence. What counts as “successful” literature search carries considerable nuance; LitBench can evaluate proposed tools at a fine-grained level.

## Introduction

Clinicians increasingly use large language model (LLM) systems to query the primary literature at the point of care.^1,2,3^ Given the potentially serious clinical consequences, relying uncritically on LLM-generated summaries is untenable. A clinically useful response should therefore go beyond a summary: it should identify the relevant papers and provide the supporting details, so that clinicians can verify the information directly. ^4^

Existing benchmarks measure several related capabilities. LitQA2 and PaperQA2 reward identifying the correct paper but do not require a quoted supporting passage.^5,6^ MIRAGE and MedRAG score free-form answers based on PubMed literature without controlling the number or composition of candidate papers.^7^ BioHopR evaluates the synthesis of preselected facts rather than the retrieval of their source papers.^8^ While individually valuable, none of these benchmarks jointly assess retrieval of the correct fact, the paper that first reported it, and the supporting passage.^9^ Furthermore, because their questionsare constructed from evidence present in the supplied or searchable evidence corpus, they do not test whether a system recognizes the absence of evidence. Four questions therefore remain: (1) whether accuracy differs between stated and inferred facts; (2) how accuracy is impacted by the number and composition of competing papers; (3) whether a system can synthesize facts from two sources; and (4) whether it declines to answer when the evidence is absent.

To address these gaps, we developed LitBench, a benchmark that evaluates evidence retrieval, fact-type, multi-paper synthesis, and appropriate abstention under controlled literature-search conditions.

## Methods

### Study Design and Overview

LitBench evaluates an end-to-end literature-search response comprising the requested fact and its source paper, and the supporting passage **(Table 1)**. We developed a custom retrieval harness for the Gemma models based on interleaving retrieval with chain-of-thought reasoning (IRCoT).^10^ Three candidate systems were evaluated across the main test conditions (T1-5): Gemma-4B + IRCoT, Gemma-12B + IRCoT, and Sonnet-5. DeepSeek-V4-Flash + IRCoT was tested only in the evidence-absent refusal conditions (T6-T7). Reporting followed TRIPOD-LLM.^11^ Figure 1 presents the LitBench report card, with credited-response rates stratified by system, fact domain, and test condition.

**Table 1.**
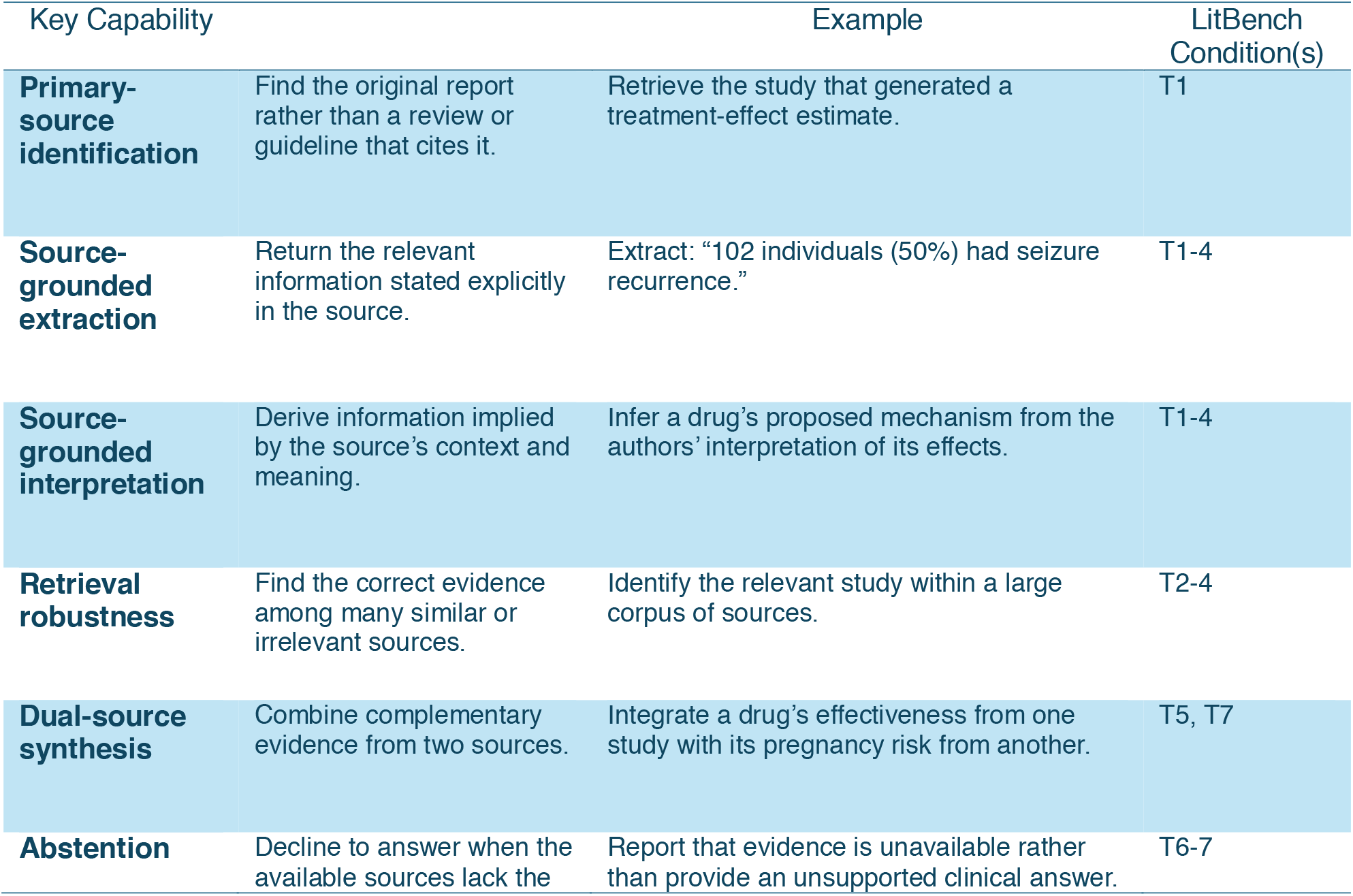
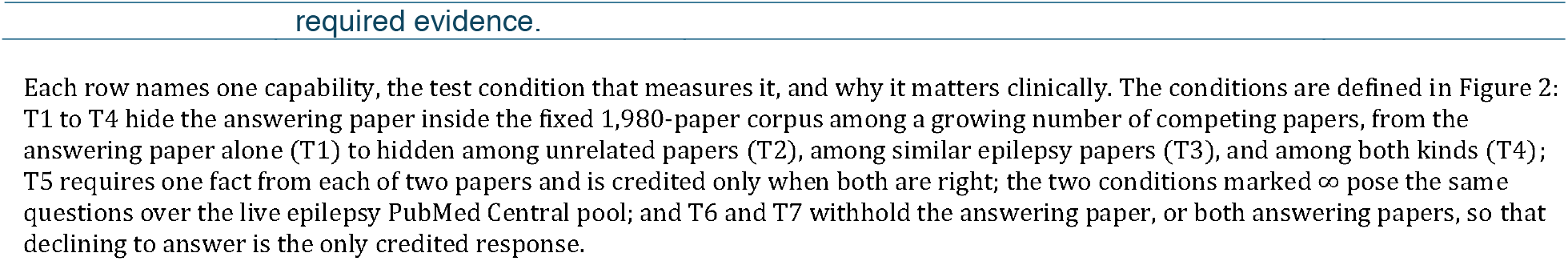
Key capabilities of a literature-retrieval system, and where LitBench tests it.

| Key Capability |  | Example | LitBench Condition(s) |
| --- | --- | --- | --- |
| **Primary-source identification** | Find the original report rather than a review or guideline that cites it. | Retrieve the study that generated a treatment-effect estimate. | T1 |
| **Source-grounded extraction** | Return the relevant information stated explicitly in the source. | Extract: “102 individuals (50%) had seizure recurrence.” | T1-4 |
| **Source-grounded interpretation** | Derive information implied by the source’s context and meaning. | Infer a drug’s proposed mechanism from the authors’ interpretation of its effects. | T1-4 |
| **Retrieval robustness** | Find the correct evidence among many similar or irrelevant sources. | Identify the relevant study within a large corpus of sources. | T2-4 |
| **Dual-source synthesis** | Combine complementary evidence from two sources. | Integrate a drug’s effectiveness from one study with its pregnancy risk from another. | T5, T7 |
| **Abstention** | Decline to answer when the available sources lack the | Report that evidence is unavailable rather than provide an unsupported clinical answer. | T6-7 |

**Figure 1.**
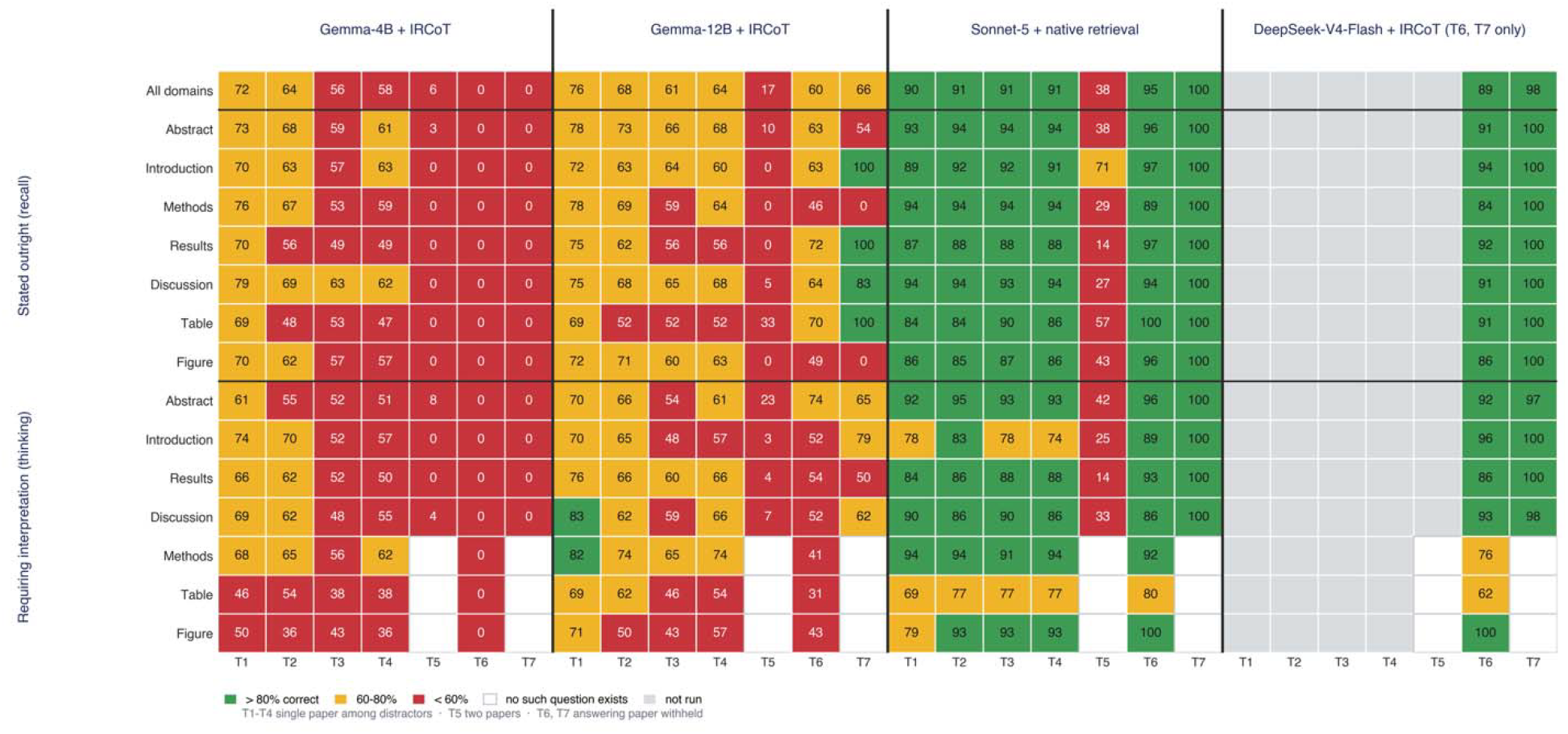
LitBench report card. Each cell shows the credited-response rate by system, fact domain, and condition. Recall facts are stated directly; interpretation facts require inference from the source. T1–T5 contain the answering evidence and are scored for the correct fact; T6–T7 withhold it and are scored for refusal. Green indicates >80%, yellow 60–80%, and red <60%; white indicates no applicable question and gray indicates not evaluated. DeepSeek-V4-Flash was evaluated only on T6–T7.

### Source Corpus and Gold-Standard Facts

The source corpus comprised 1,980 open-access papers: 1,000 epilepsy papers containing answer-bearing facts and 980 non-epilepsy decoys. Epilepsy was selected as the exemplar clinical domain because of the authors’ subject-matter expertise. A multi-agent framework performed section-specific fact extraction and reconciliation, resulting in 201,369 eligible facts (eligibility required a candidate to be useful (not boilerplate, a transition sentence, or a figure pointer), verifiable (its quote locatable in the source), originating in that paper rather than reporting a cited prior study, and not a restatement of an earlier result in the same paper; eFigure S0, eMethods S1.1). At most one fact per populated domain per paper was randomly sampled, yielding 10,254 facts for human review. A.G. reviewed all 10,254 sampled facts for correctness, source section, and classification as either explicitly stated (recall) or requiring interpretation, excluding 526 as incorrect, mis-sourced, or unusable. T.Z., working independently, reviewed 7,120 facts, covering 7,037 of the 9,728 A.G. approved, and excluded a further 256 on the same grounds (782 excluded in total). The 2,691 approved facts the second reviewer did not reach were retained, so a fact entered the gold set unless either reviewer rejected it. This yielded 9,472 gold-standard facts (eFigure S0, eMethods S1.2). Only statements from the paper that originated a claim were retained as answer-bearing evidence; subsequent restatements were not eligible for credit.

### Question Bank

Every retained fact was tagged with one of two fact types, stated outright (the source states it directly) or requiring interpretation (the reader must work it out from what the source states), crossed with its article section to give the 14 scored domains. Of the 9,472 gold facts, 8,038 (84.9%) were stated outright, 1,420 (15.0%) required interpretation, and 14 (0.1%) could not be classified. Claude Opus 4.6 generated one candidate question per fact approved on first review (n=9,728), before second-reviewer reconciliation. A.G. reviewed every question; the second reviewer read the underlying facts rather than the questions. A question had to be clinically plausible, be answerable from its supporting quote alone, correspond to a unique answering passage, 5–30 words in length, and not contain the answer. Questions that failed to meet these criteria were regenerated up to three times and excluded if they remained ineligible. To verify that interpretation-labeled questions genuinely required reasoning, we tested them using Qwen3.5-9B with reasoning disabled. The 1,207 questions the model could still answer were excluded from the interpretation question bank, although their underlying source facts were retained in the fact set. Review and reconciliation reduced the single-paper bank from 2,252 to 2,188 questions. Separately, 170 questions that require evidence from two source papers were constructed from 95 source papers (Table S1a).

### Conditions T1-T7 and How Competing Papers Were Added

For each question, N was the number of candidate papers searched (Figure 2). Identical candidate sets were used for direct system comparisons. Each system was evaluated on seven conditions (T1-7). T1 contained the answering paper alone; T2 added unrelated medical papers; T3 added similar epilepsy papers; T4 mixed both types; and T5 placed two answering papers among mixed candidates and required one fact from each. T6 and T7 used the corresponding question banks but withheld the answering paper(s) and used non-epilepsy decoys, so explicit abstention was the only correct response.^12-14^ T4·∞ and T5·∞ repeated the single- and two-paper conditions over live epilepsy PubMed Central (127,076 records; queried July 14, 2026); credit was given only if the paper that originally reported the evidence was identified.

**Figure 2.**
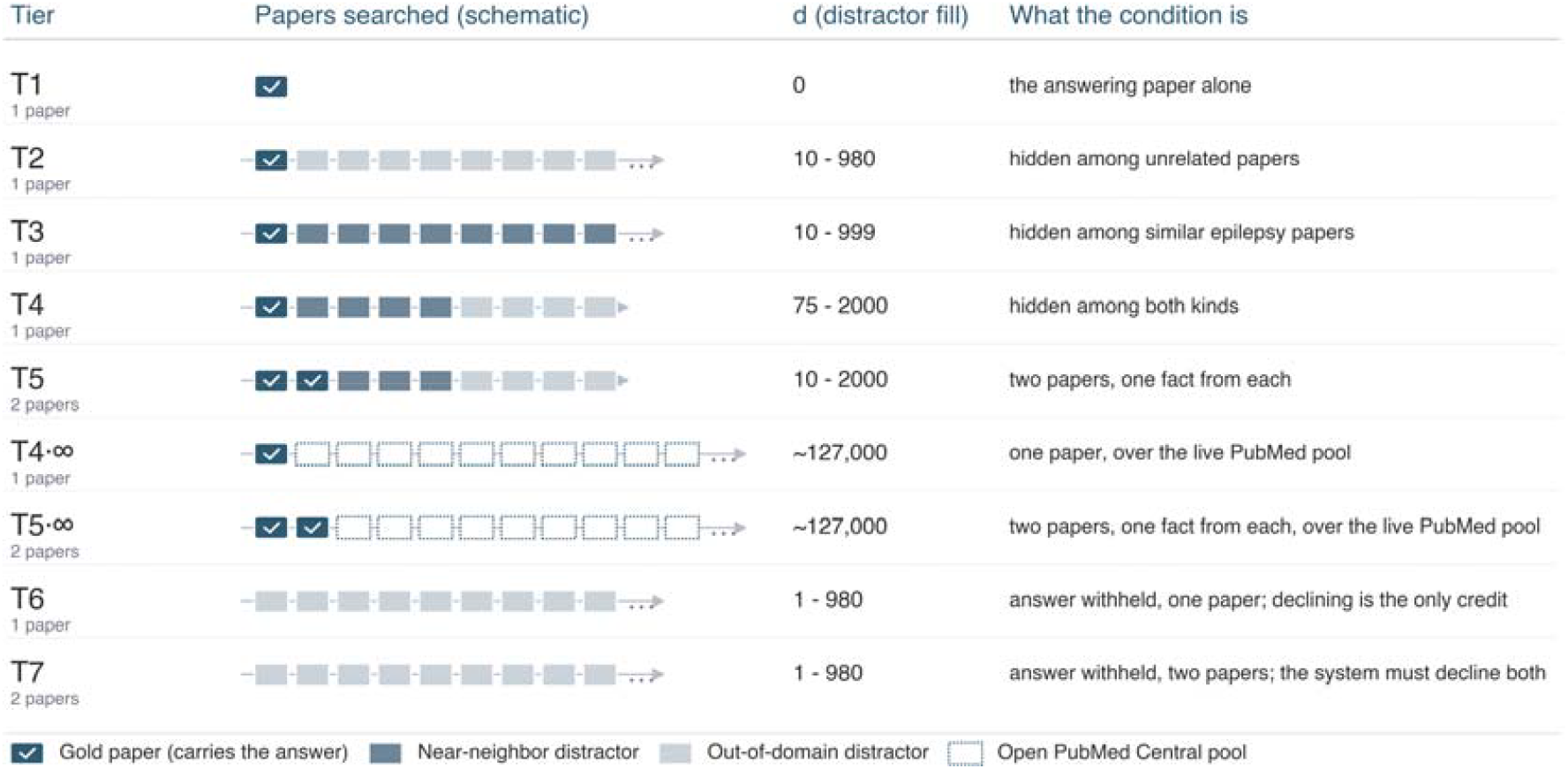
LitBench conditions and candidate-set design. N is the number of candidate papers. T1 contains one answering paper (N=1); T2 adds unrelated papers; T3 adds similar epilepsy papers; T4 mixes both; and T5 contains two answering papers and requires one fact from each. T6 and T7 mirror the single- and two-paper banks but omit the answering paper or papers, making refusal the only credited response. T4·∞ and T5·∞ use the live epilepsy PubMed Central pool. Exact candidate-set compositions at each tested N are reported in eMethods S1.5.

### Retrieval Harness for the Gemma Configurations

Both Gemma configurations shared the same IRCoT pipeline. The pipeline first retrieves eight highest-ranked passages by keyword search, then executes a reason-then-search loop under a bounded iteration cap. It returned a single verbatim quotation and used a source ledger to enforce the two-paper requirement when relevant. This pipeline was chosen as a simple, reproducible reference approach (Appendix A). The Sonnet-5-based system retrieved the evidence directly through Claude Code (i.e. no IRCoT).

### System Configurations Evaluated

Three end-to-end configurations were evaluated when evidence was present. Gemma-4B and Gemma-12B (Google; open weights; Q8_0) were served with llama.cpp on NVIDIA L40S GPUs and used the shared IRCoT harness. Sonnet-5 + native retrieval used Claude Sonnet-5 (Anthropic) through Claude Code. Because DeepSeek-V4-Flash was used as a part of the judging panel, it was only evaluated on the evidence-absent conditions (T6-7) which were scored deterministically. Gemma and DeepSeek were evaluated at six values of N on those conditions. For cost control, Sonnet-5 was evaluated on those same evidence-absent conditions on a sample rather than the full sweep: 605 single-paper items at N=50 and 390 two-paper items across N=1, 10, and 50. On the evidence-present conditions (T1-T5) Sonnet-5 was evaluated on the full question bank. Sampling parameters and complete configuration details are in eMethods and Appendix B.

### Scoring: Three-LLM Panel of Judges

Because many thousands of responses were generated that differ only in wording, we used a panel of LLM judges. Three locally hosted LLM judges (DeepSeek-V4-Flash, Qwen3.6-35B-A3B, and Llama-4-Scout) voted independently, with credit awarded by majority. For T5, both facts had to be credited independently. Judge-side reasoning was disabled, and no model judged its own output. T6–T7 were scored deterministically (no judge-panel) from the explicit refusal field. Prior work reports strong agreement between diverse LLM panels and human ratings (Cohen κ, 0.867–0.896),^15,16^ and this study included the validation described below.

### Statistical Analysis

Accuracy was defined as the proportion of credited responses in each domain-condition-N cell, averaged across the sweep at matched N. For single-paper conditions, accuracy intervals were estimated using source-clustered bootstrapping (5,000 resamples), whereas between-system differences were calculated for question-pairs using question-level bootstrapping for CI estimation. For conditions requiring two sources (T5 and T7), confidence intervals for both accuracy and between-system difference were obtained by resampling the 95 constituent source papers because many questions shared key source papers. Treating these questions as independent would fail to account for the dependence created by frequently reused hub papers (164 of the 170 two-paper pairs belonged to one connected component) (eTable S1.9). Analyses were descriptive, with no multiple-comparison corrections.

### Ethics and Reporting

LitBench used published-literature questions with no human participants or patient data. Institutional review board approval was therefore not needed. Analysis and figure-generation code and the complete manifest of the 1,980 articles used are publicly available on GitHub (https://github.com/GoldenholzLab/LitBench). To preserve the test setas uncontaminated, the exact questions are not posted publicly; complete testing materials are available from the authors on reasonable request.

## Results

### The Benchmark

The benchmark comprised 1,000 epilepsy papers and 980 non-epilepsy decoys, with 9,472 reviewed facts across the 14 fact domains (eFigure S0). These yielded 2,188 single-paper questions and 170 two-paper questions, which were evaluated under the conditions shown in Figure 2. Reported accuracy reflects credits assigned by the LLM judge panel; refusal/abstention conditions were scored deterministically (Methods).

### Single-Paper Accuracy Exceeded Two-Paper Accuracy

Single-paper accuracy exceeded two-paper accuracy for all three configurations in which the answering evidence was present (T1-T5, as opposed to the evidence-absent T6-T7) (Table 2; Figures 1 and 3). Across the fixed-corpus single-paper conditions, accuracy ranged from 55.9% to 71.9% for Gemma-4B, 61.2% to 75.5% for Gemma-12B, and 90.0% to 91.3% for Sonnet-5. Candidate-set size and composition posed distinct challenges. For both Gemma configurations, accuracy was lower with similar-paper distractors (T3) than with mixed distractors (T4); thus, T1–T4 should not be interpreted as a monotonic progression in difficulty. At T4, Sonnet-5 outperformed Gemma-12B by 27.2 percentage points (95% CI, 25.1–29.5).

**Table 2.**
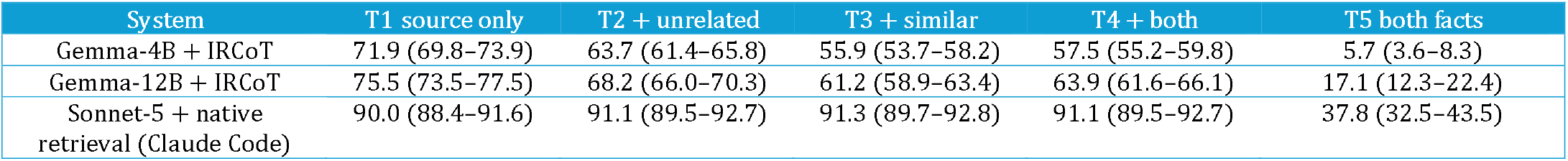
Primary semantic fact accuracy by system and condition. Values are percentages credited by majority vote of the three-model judge panel, with 95% confidence intervals. T1–T4 are single-paper conditions; T5 contains two source papers and requires both facts. Gemma-4B and Sonnet-5 had 2,188 evaluable single-paper responses per condition. Gemma-12B had 2,184; four missing responses were excluded. A separate source-and-span composite is reported in Table S4c.

| System | T1 source only | T2 + unrelated | T3 + similar | T4 + both | T5 both facts |
| --- | --- | --- | --- | --- | --- |
| Gemma-4B + IRCoT | 71.9 (69.8–73.9) | 63.7 (61.4–65.8) | 55.9 (53.7–58.2) | 57.5 (55.2–59.8) | 5.7 (3.6–8.3) |
| Gemma-12B + IRCoT | 75.5 (73.5–77.5) | 68.2 (66.0–70.3) | 61.2 (58.9–63.4) | 63.9 (61.6–66.1) | 17.1 (12.3–22.4) |
| Sonnet-5 + native retrieval (Claude Code) | 90.0 (88.4–91.6) | 91.1 (89.5–92.7) | 91.3 (89.7–92.8) | 91.1 (89.5–92.7) | 37.8 (32.5–43.5) |

**Figure 3.**
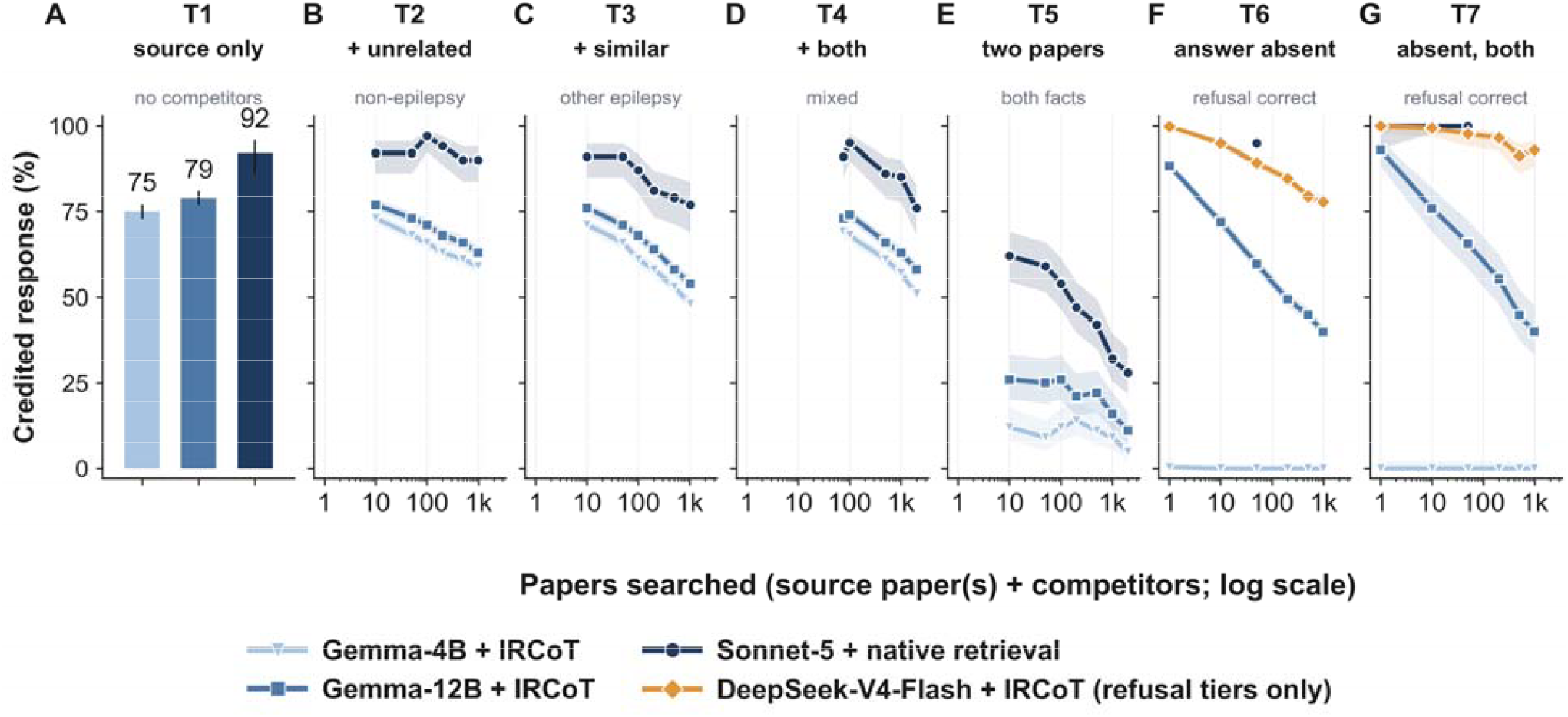
Credited responses by condition and number of candidate papers (N). Panels map directly to the benchmark conditions: A, T1 (answering paper alone); B, T2 (unrelated papers); C, T3 (similar epilepsy papers); D, T4 (mixed papers); E, T5 (two-paper evidence); F, T6 (single-paper evidence absent); and G, T7 (two-paper evidence absent). Panels A–E show semantic fact accuracy; F–G show correct refusal. B–G use logarithmic x-axes. Sonnet-5 refusal testing comprised 605 T6 cells at N=50 and 390 T7 cells across N=1, 10, and 50; Gemma and DeepSeek used six values of N. DeepSeek was evaluated only in T6-T7. Bands are 95% Wilson intervals for each N-specific cell.

Averaged across fact domains, accuracy was lower for interpretation-requiring facts compared to stated facts (by 5.5 percentage points for Gemma-4B, 3.9 for Gemma-12B, and 2.2 for Sonnet-5). The magnitude of this difference varied across configurations and fixed-corpus conditions (Figure S1).

### Two-Paper Retrieval and Synthesis Remained Difficult

On the two-paper condition (T5), accuracy was 5.7% for Gemma-4B, 17.1% for Gemma-12B, and 37.8% for Sonnet-5, compared with mean single-paper accuracy of 62.3%, 67.2%, and 90.9%, respectively (Figure S3). At least one of the two facts was credited much more often (64.5%, 76.3%, and 87.9%) than both facts together. On T5, Sonnet-5 led Gemma-12B by 20.7 percentage points (95% CI, 16.4–25.2) and Gemma-4B by 32.1 points (27.2–37.5); Gemma-12B led Gemma-4B by 11.4 points (7.4–16.0).

Retrieval logs showed that most Gemma T5 cells retrieved only one source paper. Even when both source papers were retrieved, both facts were credited in only a minority of cases (eTable S1.10; Table S6). These observations suggest both retrieval of the second source and downstream answer construction need further refinement.

Over the live PubMed Central condition (T5·∞), Sonnet-5 remained approximately 30% more accurate than Gemma-12B (Figure S1). A post hoc exploration found that searching with the full two-paper question did not retrieve both source papers, whereas separate searches for each fact did. T5·_∞_ is not directly comparable with T4·_∞_ because the former used a smaller, hand-selected question set, with each component item searched separately. The higher T5·_∞_ values therefore should not be interpreted as evidence that two-paper retrieval was easier. Live searches were performed in July 2026.

### With the Evidence Removed, Systems Differed in Their Abstention Rates

When the answering paper was withheld, Gemma-12B’s refusal rate declined as N increased: from 88.3% to 39.9% for single-paper items and from 92.9% to 40.0% for two-paper items. Gemma-4B refused only 0.07% for single-paper and 0% for two-paper (Figure 3F-G; Table S5).

For cost control, Sonnet-5’s refusal rate was evaluated at a subset of N values (N=50 for single-paper; N=1, 10, and 50 for two-paper) rather than swept across the full six-value grid used for Gemma and DeepSeek (Appendix B): it correctly refused on 574 of 605 single-paper cells at N=50 (94.9%) and all 390 two-paper cells across N=1, 10, and 50 (100%). Its substantially lower refusal rate on matched evidence-present controls suggests that it did not abstain indiscriminately. In contrast, DeepSeek-V4-Flash (evaluated across all six values of N) maintained high correct refusal rates but also refused on 80.3% of matched answer-present controls (Table S5).

### Judge Concordance

The primary panel score generally agreed with the independent source-and-verbatim-span cross-check; disagreements often involved paraphrases rather than copied supporting text. For human validation, A.G. reviewed a condition-stratified sample of 154 cells while blinded to the LLM panel verdict. Panel and clinician ratings agreed on 89.0% of the cases (Cohen κ, 0.774; 95% CI, 0.67–0.87), with the panel more likely than the clinician to award credit (Table S4b).

## Discussion

We developed LitBench, a benchmark for medical literature retrieval, and evaluated several retrieval systems. Performance generally improved with more capable models, but some conditions remained challenging for all. No system achieved uniformly high performance across all conditions. The conditions also appear to capture distinct dimensions of retrieval capability, as strong performance on one condition did not predict success in others.

The two-paper conditions were a major source of failure. These conditions required retrieval and synthesis of both source papers. The logs identify failure to retrieve the second paper as one important bottleneck. Incomplete credit after both papers were retrieved indicates a second extraction or synthesis bottleneck. The frontier system (Sonnet-5) outperformed the lower models, consistent with prior reports that general-purpose models outperform purpose-built clinical tools.^17^

No single existing framework jointly covers the capabilities clinicians need.^9^ In medicine, paper-identification benchmarks test document retrieval without requiring a supporting passage;^5,6^ free-form suites evaluate answers without a controlled candidate set;^7,18^ citation and factuality studies compare generated statements with supplied sources;^19,20^ related attribution and claim-verification tasks operate at document or abstract-sentence level;^21,22^ multi-paper benchmarks provide the component facts rather than require source retrieval;^8^ and multiple-choice suites,^23,24^ systematic-review automation,^25^ and an epilepsy knowledge-graph benchmark^26^ address other useful endpoints. Outside medicine, OpenScholar and ScholarQABench provide close comparisons,^27^ and AstaBench extends this line of work by grading answer ranking and checklist coverage;^28^ none require citing the exact passage from which a claim originates, and ScholarQABench also accepts any supporting passage, searches a large uncontrolled paper pool, calculates citation recall against the system’s own generated statements, and treats an originating report and a later restatement equivalently. Other work has evaluated pieces of this problem separately: semi-extractive question-answering benchmarks require a verbatim supporting span but supply the source papers;^29^ long-context citation benchmarks test source attribution as unrelated text is added;^30^ deep-search benchmarks control distractors but score document identifiers;^31^ and concurrent work verifies verbatim matches within supplied candidate papers.^32^

LitBench differs from all of these by jointly assessing fact type, the originating paper, the supporting passage, a controlled and graded candidate set, two-paper retrieval and synthesis, and appropriate refusal in a single clinical benchmark, and by reporting semantic fact accuracy separately from source-and-span traceability. These differences matter beyond completeness: identifying the originating report, rather than crediting any restatement, is important because later citations can propagate unsupported claims,^33^ and approximately one in six quotations across 46 studies did not support the cited claim.^34^

### Study Limitations

This study has several limitations. First, it included only open-access epilepsy papers, so the results may not apply equally to closed-access literature or other specialties. Second, the main outcome was scored by three LLM judges,^15,16^ a method also validated here by one clinician on 154 responses (Cohen κ, 0.774). Third, the systems differed in both their models and retrieval methods, making precise comparisons difficult. Fourth, the two-paper set contained only 170 questions, limiting more detailed analyses. Sonnet-5 was tested on only a sample of refusal conditions, and DeepSeek was tested only on refusal conditions. Fifth, the refusal results require caution. Gemma had no matched answer-present controls, and DeepSeek often refused even when an answer was available. The evidence-absent conditions also contained only non-epilepsy papers, so systems may have refused because the papers were off-topic rather than because they recognized that the specific answer was missing. Finally, the benchmark contains many more single-paper than two-paper questions because two-paper questions were harder to construct; this ratio does not reflect how often each question type occurs in clinical practice.

## Conclusion

LitBench measures whether an answer traces to the original report that supports it, across one paper, across two, and across the distractor grid of competing papers. It also distinguishes between recall and thinking style facts. It separated four systems without saturating on any axis: by fact domain, by the distractor grid, by two-paper synthesis, or by recognizing absent evidence. No system solved all four axes. The unsolved gap is a reminder that autonomous literature synthesis, often treated as a solved precursor to AI-assisted science, is not solved even in this bounded, single-specialty form. The longer-term goal is to use trustworthy literature retrieval in clinically relevant scenarios, both real-time at the bedside and offline for research. The longer-term clinical goal may be achieved with a single explainable system, capable of all the subtasks required by LitBench. At present we do not believe a single system is capable of this, but anticipate further research can move in this direction.

## Supporting information

full appendix

## Data Availability

All data produced are available online at https://github.com/GoldenholzLab/LitBench

## Acknowledgments

Large language models appear in this work in two distinct roles. As study systems they are the object of the evaluation and are specified in the Methods and in eMethods S1.6 to S1.8; this includes Claude Opus 4.6, which drafted the candidate questions. Separately, in preparing this manuscript, ChatGPT (GPT-5.6 Sol, OpenAI) was used solely for grammar, language polishing, and formatting. No language model produced or altered any result, number, figure or reference. The authors reviewed and edited all AI-assisted text, take full responsibility for the content of this manuscript, and list no language model as an author.

## Author contributions

AG: conception, data collection, analysis, drafting manuscript. TZ: data collection, editing. MBW: editing. DG: conception, oversight, editing.

## References

1. Omar M, Soffer S, Agbareia R, et al. Sociodemographic biases in medical decision making by large language models. Nat Med. 2025;31(6):1873–1881. doi:10.1038/s41591-025-03626-6

2. Thirunavukarasu AJ, Ting DSJ, Elangovan K, Gutierrez L, Tan TF, Ting DSW. Large language models in medicine. Nat Med. 2023;29(8):1930–1940. doi:10.1038/s41591-023-02448-8

3. Zakka C, Shad R, Chaurasia A, Dalal AR, Kim JL, Moor M, et al. Almanac — retrieval-augmented language models for clinical medicine. NEJM AI. 2024;1(2). doi:10.1056/AIoa2300068

4. Goodman KE, Yi PH, Morgan DJ. AI-generated clinical summaries require more than accuracy. JAMA. 2024;331(8):637–638. doi:10.1001/jama.2024.0555

5. Laurent JM, Janizek JD, Ruzo M, et al. LAB-Bench: measuring capabilities of language models for biology research. arXiv preprint arXiv:2407.10362. 2024.

6. Skarlinski MD, Cox S, Laurent JM, et al. Language agents achieve superhuman synthesis of scientific knowledge. arXiv preprint arXiv:2409.13740. 2024.

7. Xiong G, Jin Q, Lu Z, Zhang A. Benchmarking retrieval-augmented generation for medicine. In: Findings of the Association for Computational Linguistics: ACL 2024:6233–6251. doi:10.18653/v1/2024.findings-acl.372

8. Kim Y, Abdulle Y, Wu H. BioHopR: a benchmark for multi-hop, multi-answer reasoning in the biomedical domain. In: Findings of the Association for Computational Linguistics: ACL 2025:12894–12908. doi:10.18653/v1/2025.findings-acl.668

9. Bedi S, Liu Y, Orr-Ewing L, Dash D, Koyejo S, Callahan A, et al. Testing and evaluation of health care applications of large language models: a systematic review. JAMA. 2025;333(4):319–328. doi:10.1001/jama.2024.21700

10. Trivedi H, Balasubramanian N, Khot T, Sabharwal A. Interleaving retrieval with chain-of-thought reasoning for knowledge-intensive multi-step questions. In: Proceedings of the 61st Annual Meeting of the Association for Computational Linguistics (Volume 1: Long Papers). 2023:10014–10037. doi:10.18653/v1/2023.acl-long.557

11. Gallifant J, Afshar M, Ameen S, et al. The TRIPOD-LLM reporting guideline for studies using large language models. Nat Med. 2025;31(1):60–69. doi:10.1038/s41591-024-03425-5

12. Rajpurkar P, Jia R, Liang P. Know what you don’t know: unanswerable questions for SQuAD. In: Proceedings of the 56th Annual Meeting of the Association for Computational Linguistics (Volume 2: Short Papers). 2018:784–789. doi:10.18653/v1/p18-2124

13. Chen J, Lin H, Han X, Sun L. Benchmarking large language models in retrieval-augmented generation. Proceedings of the AAAI Conference on Artificial Intelligence. 2024;38(16):17754–17762. doi:10.1609/aaai.v38i16.29728

14. Kamath A, Jia R, Liang P. Selective question answering under domain shift. In: Proceedings of the 58th Annual Meeting of the Association for Computational Linguistics. 2020:5684–5696. doi:10.18653/v1/2020.acl-main.503

15. Verga P, Hofstatter S, Althammer S, et al. Replacing judges with juries: evaluating LLM generations with a panel of diverse models. arXiv preprint arXiv:2404.18796. 2024.

16. Zheng L, Chiang WL, Sheng Y, et al. Judging LLM-as-a-judge with MT-Bench and Chatbot Arena. In: Advances in Neural Information Processing Systems 36: Datasets and Benchmarks Track. 2023:46595–46623. doi:10.52202/075280-2020

17. Vishwanath PR, et al. General-purpose large language models outperform specialized clinical artificial intelligence tools. Nat Med. 2026. doi:10.1038/s41591-026-04431-5

18. Bedi S, Cui H, Fuentes M, et al. Holistic evaluation of large language models for medical tasks with MedHELM. Nat Med. 2026;32(3):943–951. doi:10.1038/s41591-025-04151-2

19. Wang X, Tan M, Jin Q, et al. MedCite: can language models generate verifiable text for medicine? In: Findings of the Association for Computational Linguistics: ACL 2025:18891–18913. doi:10.18653/v1/2025.findings-acl.967

20. Min S, Krishna K, Lyu X, et al. FActScore: fine-grained atomic evaluation of factual precision in long-form text generation. In: Proceedings of the 2023 Conference on Empirical Methods in Natural Language Processing. 2023:12076–12100. doi:10.18653/v1/2023.emnlp-main.741

21. Gupta D, Demner-Fushman D, Hersh W, Bedrick S, Roberts K. Overview of the TREC 2024 Biomedical Generative Retrieval (BioGen) Track. In: Proceedings of the Thirty-Third Text REtrieval Conference (TREC 2024). National Institute of Standards and Technology; 2024. doi:10.6028/nist.sp.1329.overview.biogen-coordinators

22. Wadden D, Lo K, Kuehl B, et al. SciFact-Open: towards open-domain scientific claim verification. In: Findings of the Association for Computational Linguistics: EMNLP 2022. 2022:4719–4734. doi:10.18653/v1/2022.findings-emnlp.347

23. Jin Q, Dhingra B, Liu Z, Cohen WW, Lu X. PubMedQA: a dataset for biomedical research question answering. In: Proceedings of the 2019 Conference on Empirical Methods in Natural Language Processing and the 9th International Joint Conference on Natural Language Processing (EMNLP-IJCNLP). 2019:2567–2577. doi:10.18653/v1/D19-1259

24. Tsatsaronis G, Balikas G, Malakasiotis P, et al. An overview of the BioASQ large-scale biomedical semantic indexing and question answering competition. BMC Bioinformatics. 2015;16:138. doi:10.1186/s12859-015-0564-6

25. Wang Z, Cao L, Danek B, Jin Q, Lu Z, Sun J. Accelerating clinical evidence synthesis with large language models. npj Digit Med. 2025;8(1):509. doi:10.1038/s41746-025-01840-7

26. Dai Y, et al. EpiGraph: building generalists for evidence-intensive epilepsy reasoning in the wild. arXiv. Preprint posted online May 2026. arXiv:2605.09505

27. Asai A, He J, Shao R, et al. Synthesizing scientific literature with retrieval-augmented language models. Nature. 2026;650(8103):857–863. doi:10.1038/s41586-025-10072-4

28. Bragg J, et al. AstaBench: rigorous benchmarking of AI agents with a scientific research suite. In: International Conference on Learning Representations (ICLR 2026). 2026. arXiv:2510.21652

29. Schuster T, Lelkes A, Sun H, et al. SEMQA: semi-extractive multi-source question answering. In: Proceedings of the 2024 Conference of the North American Chapter of the Association for Computational Linguistics: Human Language Technologies (Volume 1: Long Papers). 2024:1363–1381. doi:10.18653/v1/2024.naacl-long.74

30. Tang Z, Zhou K, Li J, Ji B, Hou J, Zhang M. L-CiteEval: a suite for evaluating fidelity of long-context models. In: Proceedings of the 63rd Annual Meeting of the Association for Computational Linguistics (Volume 1: Long Papers). 2025:5254–5277. doi:10.18653/v1/2025.acl-long.263

31. Chen Z, Ma X, Zhuang S, et al. BrowseComp-Plus: a fair and disentangled evaluation benchmark for deep search agents. In: Proceedings of the 64th Annual Meeting of the Association for Computational Linguistics (Volume 1: Long Papers). 2026:22349–22370. doi:10.18653/v1/2026.acl-long.1023

32. Imran M, Solanky V. ResearchQA: evaluating scholarly question answering with verbatim citation accuracy. arXiv. Preprint posted online July 13, 2026. arXiv:2607.11074

33. Greenberg SA. How citation distortions create unfounded authority: analysis of a citation network. BMJ. 2009;339:b2680. doi:10.1136/bmj.b2680

34. Baethge C, Jergas H. Systematic review and meta-analysis of quotation inaccuracy in medicine.Res Integr Peer Rev. 2025;10(1):13. doi:10.1186/s41073-025-00173-z

