## Supplementary material for "LitBench: A Benchmark for Retrieval-Grounded Multi-Paper Evidence Synthesis in Epilepsy": full appendix

This Supplementary Information accompanies the main manuscript. It contains the rationale for the fixed reference retrieval harness (Appendix A) and a Sonnet-5 refusal-tier sample (Appendix B), expanded Methods for corpus construction, question generation, retrieval, model serving, and scoring (S1), the supplementary tables and figures referenced in the main text (S2, S3), the judge-validation protocol (S4), and code and data availability (S5).

### Appendix A. Selection of the Retrieval Harness for the Gemma Configurations

Both Gemma configurations shared one retrieval harness, interleaving retrieval with chain-of-thought reasoning (IRCoT). This appendix explains why. It is secondary to the benchmark itself and does not report a claim of the main study. The Sonnet-5-based configuration did not use this harness: it performed its own retrieval, so the configurations are compared as complete end-to-end systems rather than as models sharing one harness.

#### A.1 Selection criterion

The harness for the Gemma configurations had to run on a local open-weight model, so these configurations were reproducible without a proprietary model in the retrieval loop, and it had to be simple enough that an independent reader could re-run it from the released artifacts. Simplicity was weighted above raw score on the screen below: a harness that scored higher but added inference cost or an extra processing stage was not preferred.

#### A.2 Harnesses evaluated

Candidate harnesses were compared on the Gemma-4B model on the two-paper tier, two papers with one fact from each, credited only when both are right (T5), at matched values of N, the number of papers searched, across the full sweep (Figure A1). The candidates spanned a one-shot retrieval-augmented generation head, the iterative reason-then-retrieve pipeline (IRCoT), decomposed per-fact retrieval, self-consistency voting over multiple sampled answers, and an agentic verify-and-refine loop, alongside the Sonnet-5-based system as a reference point. The full screen is available from the authors on reasonable request (S5).

#### A.3 Why IRCoT

IRCoT was not the highest-scoring harness in this screen. Self-consistency voting and decomposed per-fact retrieval both scored higher on two-paper synthesis (Figure A1): averaged across the full sweep of N, self-consistency voting reached about 15.9% against about 10.3% for IRCoT, and the gap was widest at the smallest N, 24.7% versus 12.4% at d = 10. Neither was adopted, because each adds machinery that would make the local configurations harder to reproduce and to compare on equal terms: self-consistency voting multiplies inference cost by sampling and voting across several answers, and decomposed per-fact retrieval adds a separate decomposition stage. IRCoT was chosen instead as the simplest harness among those screened, using only a bounded reason-then-retrieve loop with a single search per reasoning step, which keeps it reproducible and portable to the open corpus. The Gemma configurations are therefore best read as fixed, reproducible illustrative configurations, not as cost-optimized competitors for the highest score in this screen.

The agentic verify-and-refine loop was the one candidate that scored below plain retrieval (simple bm25) in this screen, because its verify-and-refine step pruned correct facts before emission: the loop emitted its facts reliably, so the loss came from the control logic discarding correct content rather than from an inability to retrieve it. Naive agentic heads and deep-research retrieval variants either floored near zero or over-filtered, and are reported with the full negative-result set in the harness panel (S3). Added agentic machinery therefore did not improve two-paper synthesis on Gemma-4B and sometimes hurt it.

Figure A1 (S3) shows this harness screen across the full sweep of N; Figure A2 (S3) shows the IRCoT pipeline itself.

### Appendix B. Sonnet-5 sampling on the refusal tiers

Sonnet-5 was evaluated on the two refusal tiers, answer withheld with one paper (T6) and with two (T7), as a bounded sample rather than across the full grid, and is reported here rather than in Table S5. Because no gold paper is present, a system cannot terminate its search on a hit and must search every paper, so a refusal item is substantially more expensive than the equivalent item on T1 to T5. Available compute, not a stopping rule, set the size of the sample. On the cells completed, Sonnet-5 declined on every two-paper cell evaluated (390 of 390, at d = 1, 10 and 50) and on 94.9% of single-paper cells (574 of 605, at d = 50).

A matched control arm quantifies what the refusal option costs. The same instruction was run with the gold paper present, so that permission to decline was the only thing differing from an ordinary retrieval item. On single-paper controls Sonnet-5 wrongly declined 6.7% (N = 1), 5.0% (N = 10), and 6.8% (N = 50) of items, a floor that stays flat rather than rising with N. On two-paper controls it wrongly declined 13.3% (N = 10) and 21.7% (N = 50); this rise tracks the harness's falling chance of retrieving and verifying both gold papers as more distractors are added, not a growing tendency to refuse. Repeat runs of the same control cells agreed on 29 of 33 occasions, and every disagreement fell on a control rather than an unanswerable item, so this floor carries run-to-run variation that the near-ceiling refusal rates do not.

These runs used the retrieval harnesses published for the single-paper tiers (T1 to T4) and the two-paper tier (T5) respectively, not the refusal-tier harness of Table S5, and are therefore not row-comparable with the Gemma configurations.

### S1. Supplementary Methods

#### S1.1 Corpus construction and the gold-fact taxonomy

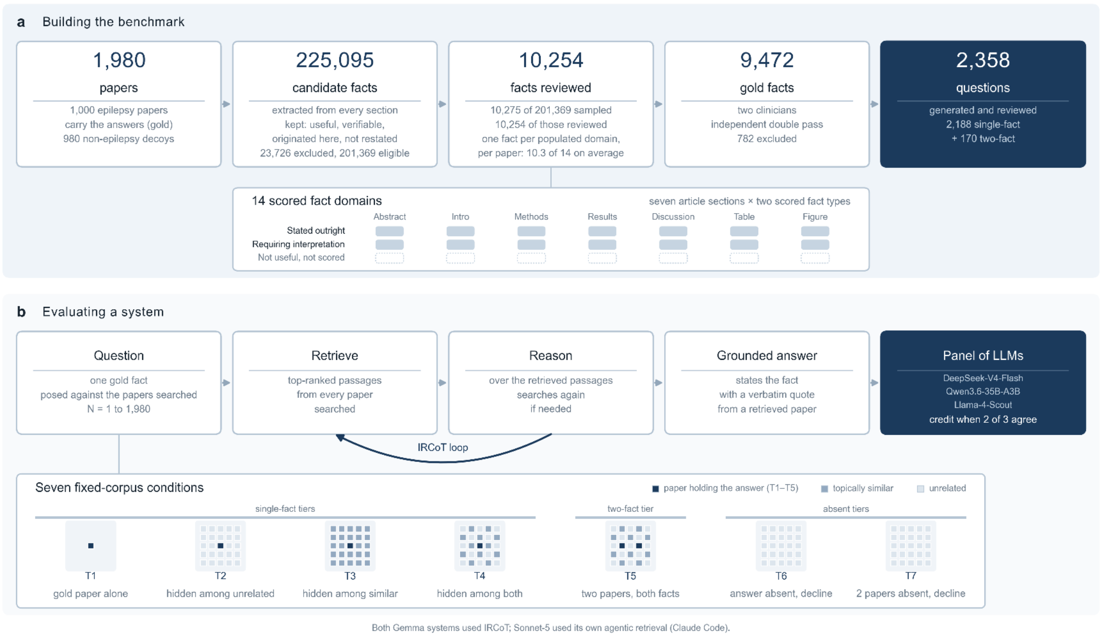


***eFigure S0. Building and testing LitBench.*** *(a) Benchmark construction. The corpus is 1,980 open-access papers, of which only 1,000 are about epilepsy; the other 980 are non-epilepsy papers that carry no answer and serve only as competing papers to search through. A gold fact is a claim taken from one of the 1,000 epilepsy papers, together with the sentence that states it, which the benchmark treats as the correct answer. Those 1,000 papers were mapped by section, and candidate facts were extracted by a multi-agent pipeline: seven section-specific language-model agents run in parallel, one per article section, followed by a reconciling agent that reads the full paper and drops, retags, or adds facts. Inclusion criteria removed 23,726 candidates, leaving 201,369 eligible, from which 10,275 were sampled for manual review, one fact per populated domain within each paper across the 1,000 papers; 10,254 of those sampled facts were reviewed. No paper populates all 14 domains, so the ceiling of 14,000 is never approached: the realized mean is 10.3 domains per paper. Of the 10,254, 526 were rejected on first review and a further 256 on second-reviewer reconciliation, leaving 9,472 gold facts. Questions were drafted from the 9,728 facts approved on first review, before that second-reviewer exclusion; a question entered the evaluation bank only if it survived three further checks, each applied to every draft: it had to be answerable from its own quote, no other passage in the corpus could answer it, and its wording could not give the quote away. A fourth check then applied to the drafts whose gold fact required interpretation, and to those only: a small model shown only the relevant section could not answer it without reasoning. Most drafts failed one of these, which is why the bank is far smaller than the fact set: 9,728 drafts fell to 8,810 after the answerability check, then to 7,672 after the second pass of that same manual answerability review, then to 5,612 after the uniqueness check and 3,459 after the give-away check. The reasoning check, which ran over the interpretation-requiring drafts alone, brought that to 2,252. Reconciling the two clinicians' fact ratings then left 2,188 single-fact questions over 835 papers, plus 170 two-*

*fact questions. The gold facts whose questions were dropped remain in the released fact set; they were never posed to a system. eMethods S1.2 gives the counts at each check, including both passes of the answerability review. Every fact is tagged by article section and fact type, giving the 14 scored domains (seven sections × two fact types) shown in the strip. (b) Evaluating a system. Each question is posed against N papers, the set the system must search through: the paper reporting the answer, where the answer is present, together with the competing papers added around it. The system retrieves passages, reasons over them, and returns a fact with a verbatim supporting quote. The Gemma-based systems interleave retrieval and reasoning with chain-of-thought (the IRCoT loop); the Sonnet-5-based system used its own agentic retrieval instead. Credit is decided by a panel of three LLM judges, each casting a binary same-fact vote, with credit awarded when two of the three agree. This figure shows the seven conditions built from the fixed 1,980-paper corpus: the answering paper alone (T1), hidden among unrelated papers (T2), among similar epilepsy papers (T3), among both kinds (T4), and two papers with one fact from each (T5). Two further conditions withhold the answering paper: answer withheld, one paper (T6), and answer withheld, two papers (T7), so no correct answer is present and an explicit refusal is the only credited response. The study ran nine conditions in all; the remaining two (T4·∞ and T5·∞) replace the fixed corpus with the live epilepsy PubMed Central pool and are shown in Figure 2 of the main text. Full construction detail follows (eMethods S1.1 to S1.5).*

The 1,000 gold papers (each carrying at least one scored fact) were drawn from PubMed through the NCBI E-utilities interface, queried once per study-type by journal bucket over open-access epilepsy and neurology journals published between 2016 and 2026 and de-duplicated by digital object identifier; study-type buckets (randomized trial, cohort, meta-analysis, guideline, case report, and genome-wide association study) were filled toward target quotas, and the query was deterministic and seed-pinned so the same paper set returns on rebuild. The remaining 980 corpus papers are distractors that carry no scored fact and serve only as competing papers. The gold set drew chiefly from Frontiers in Neurology and Epilepsia Open (each about 38%), then Scientific Reports (about 14%), with the remainder from PLoS ONE and BMC Neurology. It contains no papers from the flagship Epilepsia or Seizure journals; this is a corpus-build artifact, not an open-access limitation; a bug in the automated-download code failed to fetch papers that were not indexed in the PubMed Central open-access mirror and needed one of the other three sources, even though downloadable open-access Epilepsia papers existed at build time. This gap is disclosed as a limitation in the main text. For each paper, full text was obtained by walking four open-access sources in priority order (the PubMed Central open-access mirror, Unpaywall, an institutional proxy, and direct resolution of the digital object identifier) and stopping at the first valid PDF, with PubMed Central supplying about 85%. Each paper was then mapped once by a language model under a strict JSON schema into its constituent sections with their page ranges, its table and figure captions, its paper type, and its studied-drug names, which provided the per-section slices the extraction agents consumed.

Facts were extracted from open-access epilepsy papers by seven section-specific language-model agents run in parallel, one per article section (abstract, introduction, methods, results, discussion, table, and figure). A further agent in the same framework classified every candidate as the originating statement of its claim or as a propagator restating a claim originated elsewhere. The corpus was searched for verbatim occurrences of the candidate's quote, and each occurrence was classified as an originator or a propagator statement. Candidates whose claim was originated by more than one corpus paper were dropped, so that strict citation scoring remained fair. This is an extraction-stage step, not part of the manual review that follows. Each agent saw only its own section and a section-tuned prompt and emitted one candidate fact per atomic claim, each with a required verbatim supporting quote, its source section, and its source page; the agents extracted greedily and left all quality and usefulness filtering to the downstream checks. This produced 225,095 candidate facts before filtering. Each candidate fact passed five sequential validation checks: an original-data check keeping only facts that report this paper's own data rather than a cited prior study (removing citation leakage), a usefulness check removing transition sentences, framing prose, and figure pointers, a fact-type check that labels each fact as stated outright or requiring interpretation and stamps its section-by-fact-type domain (a labeling step with no drops), an anti-rehash check removing Discussion sentences that merely restate earlier results, and a quote-grounding check failing facts whose verbatim supporting quote cannot be located in the source. Surviving facts were labeled under a 14-domain taxonomy formed by crossing seven article sections with two fact types, stated outright (a fact the source states directly) and requiring interpretation (a fact the reader must work out from what the source states); the exact seven-section list is recorded with the corpus artifact. Three interpretation-requiring domains (methods, table, and figure) carry no two-paper questions by construction, so they appear as structurally not-applicable cells in the two-paper tiers, T5 and its open-corpus counterpart.

Beyond these automated quality checks, gold-set construction proceeded through four further stages: manual review of every candidate fact by the first reviewer and of a subset by an independent second reviewer (S1.2), a fake-reasoning filter that relabeled rather than discarded interpretation-requiring facts, a length filter restricting retained single-paper items to a 5-to-30-word range, and a manual review of the generated questions by the first reviewer alone. These stages are detailed below (S1.2).

A seed-42 stratified sample drew 10,275 facts from the filtered pool for manual review by two independent reviewers; 10,254 were reviewed in the first reviewer's pass (21 were not processed), of which 9,728 were approved and 526 were rejected on review. The remainder of the filtered pool was not sampled and is described as excluded, not rejected.

#### S1.2 Two-reviewer reconciliation of the gold set

The first reviewer manually reviewed all 10,254 sampled candidate facts. The second reviewer, working independently, reviewed 7,120 of them, covering 7,037 of the 9,728 facts the first reviewer approved; the remaining 2,691 approved facts were not reached in the second pass. The two reviews were then reconciled through a three-signal merge: a fact was retained in gold only if it was approved by the first reviewer and not rejected or flagged as unusable by the second reviewer, so a fact the second reviewer never reached was retained; and facts flagged by a fake-reasoning filter were relabeled from requiring interpretation to stated outright rather than dropped. That filter re-answered every fact labeled as requiring interpretation with a separate small model (Qwen3.5-9B), reasoning disabled and shown only the fact's own section rather than the full paper so the answer could not be located elsewhere, and a same-or-different judge compared the answer to the gold claim; a fact recoverable without reasoning was relabeled as stated outright and was kept, never dropped, unless the second reviewer read it as requiring interpretation and restored the label. The same labels were applied differently to the question bank: a question whose gold fact was flagged was removed from the bank rather than relabeled (1,207 of 3,459 removed), because a question a small model can answer without reasoning does not test what the interpretation-requiring domains are for. The fact itself stays in the gold set with its demoted label. The merge reduced the approved pool from 9,728 to 9,472 facts. The 256 facts the second reviewer excluded left the gold set. The single-paper evaluation bank fell with it, from 2,252 to 2,188 questions. The second reviewer read facts, not questions; the questions were reviewed by the first reviewer alone. A question left the bank when the fact behind it was excluded, and 64 questions left on that basis (consort/build_reconciled_map.py). A further 309 questions kept their place but carry a gold fact relabeled from requiring interpretation to stated outright. A length filter, applied when single-paper questions were generated from approved facts, separately restricted retained items to a 5-to-30-word range (Table S1a). Across both review passes, 782 of the 10,254 first-pass-reviewed facts were excluded from gold (526 rejected on first review, 256 excluded on second-reviewer reconciliation). The 170 manually reviewed two-paper questions were not modified by the reconciliation, because the single-paper fake-reasoning filter does not apply to validated two-fact syntheses.

**Table S1a. Corpus-to-evaluation item flow.** Counts trace to the release manifest (release_manifest.json) and the reconciliation record (reconciled_query_map.json); the two-reviewer merge of the gold facts is described above. Every question-construction count is a distinct (paper, fact) key after Unicode NFC normalization of the paper identifier; the files underlying those checks store a minority of accented paper identifiers in both normalization forms, so raw row counts run slightly higher. At every fact-construction stage, entering = retained + excluded/unprocessed. Fake-reasoning-flagged facts were relabeled (requiring interpretation to stated outright) and stay in the gold fact set, so that relabeling is not an exclusion at the fact-reconciliation row above. The corresponding draft questions, by contrast, were dropped from the bank rather than relabeled (the fake-reasoning-filter row below, and the paragraph following the table).

| Construction stage | Entering | Retained | Excluded / unprocessed |
|---|---:|---:|---:|
| Corpus papers (gold + distractor) | — | 1,980 (1,000 + 980) | — |
| Facts extracted | — | 225,095 | — |
| Eligible after quality checks | 225,095 | 201,369 | 23,726 excluded (failed a quality check) |
| Sampled for review (seed 42) | 201,369 | 10,275 | 191,094 not sampled |
| Reviewed, first pass | 10,275 | 10,254 | 21 unprocessed |
| First-pass approved (reviewer 1) | 10,254 | 9,728 | 526 rejected |
| Reconciled gold (reviewer 1 and reviewer 2; fake-reasoning relabeled) | 9,728 | 9,472 | 256 excluded by second reviewer |

| Construction stage | Entering | Retained | Excluded / unprocessed |
|---|---|---|---|
| Candidate questions drafted | — | 9,728 | — |
| Answerability review, first pass (manual clinician review) | 9,728 | 8,810 | 918 not answerable |
| Answerability review, second pass (manual clinician review)* | 8,810 | 7,672 | 1,138 not answerable from the gold fact |
| No other corpus passage answers them | 7,672 | 5,612 | 2,060 not unique |
| Wording does not give the quote away† | 5,612 | 3,459 | 2,153 shortcut or ungrounded |
| Not answerable without reasoning by a small model (fake-reasoning filter) | 3,459 | 2,252 | 1,207 answerable |
| Single-paper evaluation bank, after the gold-fact reconciliation | 2,252 | 2,188 | 64 excluded |
| Two-paper synthesis questions | 178 | 170 | 8 excluded |

* This row is the second pass of the same manual answerability review recorded in the row above, not a separate automated filter, which is why no standalone script implements it: the first reviewer (A.G.) read each surviving draft against its gold fact and removed those the fact could not answer, including drafts whose rewrite-and-recheck attempts still failed (S1.3). Across both passes the review removed 2,056 of the 9,728 drafts on this single criterion. The removed rows carry its signature: they are disproportionately short, single-value facts whose question has no answerable referent in the fact (mean quote length 65 characters, vs 145 for retained rows; removal rate 30.5% for table-sourced drafts, 19.4% for methods-sourced and 16.9% for figure-sourced, against 2.5% for discussion-sourced; 23% of removed rows are table-sourced vs 8% of retained). A representative removed item pairs the gold quote “5” with the question “What

numeric value was reported in the table entry?", which that fact cannot answer. The before-and-after row manifests (litbench/qgen/g2_checked_final.parquet and litbench/qgen/g2_checked_pruned.parquet) are available from the authors on reasonable request, so that the selection can be audited independently.

† The wording-check drop log (g5_filtered.parquet, g5_dropped.parquet) records a reason for 2,155 distinct keys: 1,970 whose answer quote could not be located in the paper body, 169 dropped on question-quote proximity and 16 on lexical overlap. Two of those keys, both from accented-author papers stored in both Unicode normalization forms, also appear in the survivor file and are counted here as retained, so this row excludes 2,153.

**Why the evaluation bank is much smaller than the gold fact set. A question was drafted from every approved gold fact, but a draft entered the bank only if it survived three further checks, each present to stop a question being answerable without the retrieval the benchmark is testing: it had to be answerable from its own quote, which the clinician co-author (A.G.) checked manually in two passes, removing 2,056 drafts in all (Table S1a); no other passage in the corpus could answer it, adjudicated by BM25 retrieval scored with Gemini 2.5 Pro; and its wording could not overlap or give away the quote, with the quote locatable in the paper body, checked by the wording-overlap screen and its rewrite loop. A fourth check then ran over the drafts whose gold fact required interpretation, and over those only: a small model (Qwen3.5-9B) shown just that fact's own section with reasoning disabled could not recover the answer. Most drafts failed at least one of these. The surviving 2,252 single-fact questions cover 842 gold papers, exactly one question per (paper, domain) pair; reconciling the two clinicians' ratings of the underlying facts then took them to 2,188 over 835 papers. Gold facts whose draft questions were dropped remain in the released fact set and were never posed to a system, so no question was run and then discarded on its score. Per-domain counts are uneven by construction, from 448 abstract stated-outright questions to 18 table interpretation-requiring questions, because the checks bite hardest where a paper's section is short.**

**A separate construction-time count.** litbench/qgen/rebuild_final.parquet records 9,438 single-fact questions from a later, independent one-question-per-fact regeneration over the 9,728 pre-reconciliation fact pool. It is **not** the parent set of the evaluation bank: 13 of the bank's 2,252 (paper, fact) keys do not appear in it. It is reported here only because earlier drafts of this supplement cited it, and it is not used in any result.

The single-paper evaluation bank is the curated subset of the generated pool that was run on the benchmark and carried through the gold-fact reconciliation. Per-system single-paper runs were 2,252 (Gemma-4B), 2,252 (Gemma-12B), and 2,188 (Sonnet-5); the reconciled set common to all three systems, used for the matched between-system contrasts, is the full bank of 2,188, so no single-paper item is excluded from any between-system comparison. This required a completion step for one system. The Sonnet-5 native-retrieval run was originally generated as a fixed 2,059-item set before the gold-fact reconciliation above was finalized, leaving a 129-item difference (2,188 − 2,059) attributable entirely to run coverage rather than to any outcome-dependent exclusion: no item was ever run and then dropped for a low or incorrect score. Those 129 items were the questions used in an earlier 132-question pilot sweep that characterized accuracy across the full range of N; the subsequent full-bank run took its work list from the items the pilot had not covered, so the 129 were never submitted to it and were therefore absent from the full-bank run and from every verdict file derived from it (consort/common_subset_report.py). Those 129 items were subsequently generated and scored for Sonnet-5 under conditions matched to the original run (consort/build_sonnet_129_cells.py, consort/merge_gap129_verdicts.py), taking that system to the full 2,188. Their composition (Table S1b) is close to uniform across the seven article sections and both fact types, consistent with a coverage gap rather than a selective one; adding them changed every reported single-paper cell by 0.4 percentage points or less. All three systems were run on the full 170-question two-paper set.

**Table S1b. Domain and fact-type distribution of the 129 single-paper bank items added to the Sonnet-5 run to complete the bank.** Computed from the reconciled bank (reconciled_query_map.json) restricted to the items that were absent from the original 2,059-item Sonnet-5 run (consort/common_subset_report.py; consort/out/common_subset_report.json). Section and fact-type labels are post-reconciliation (fake-reasoning items relabeled from Requiring interpretation to Stated outright).

| Domain | Added (of 129) |
|---|---|
| Introduction | 20 |
| Discussion | 19 |
| Table | 19 |
| Methods | 18 |
| Abstract | 18 |
| Results | 18 |
| Figure | 17 |

| Fact type | Added (of 129) |
|---|---|
| Stated outright | 86 |
| Requiring interpretation | 43 |

#### S1.3 Single-paper question generation

Single-paper questions were produced by a multi-stage pipeline. A single language model (Claude Opus 4.6) generated one clinical-style question from each approved fact’s verbatim quote. The first reviewer (A.G.) then manually checked each question against its gold quote for answerability, with up to three rewrite-and-recheck attempts; the second reviewer read facts only. Two further automated stages followed: a uniqueness check in which a language model was shown the question alongside passages retrieved from the most similar non-source papers in the corpus and asked whether any of them could also answer it, dropping ambiguous items; and a tagging stage assigning the 14-domain label.

#### S1.4 Two-paper question construction

Two-paper questions were composed from pairs of single-paper facts drawn from two different gold papers through the following stages: pairwise synergy detection (retain pairs for which a clinically natural question requires both facts); composition of the question text with a deterministic leak check ensuring the question did not contain either answer; the same answerability, cross-corpus uniqueness, and tagging checks applied to single-paper questions, adapted to require both facts; an ablation check that rejected any question answerable from either fact alone or from general clinical knowledge; and a manual review of every surviving question. Building a two-paper question was a deliberate authored task, not an automatic combination of two single-paper questions: each of the 170 questions used for the main results was manually reviewed and fuses two facts from two distinct papers into one clinically coherent question that requires both facts to answer, not reducible to two separate single-paper lookups; each is mapped to the two domains of its constituent facts, and credit required facts from two distinct papers.

#### S1.5 How competing papers were added, and the open-corpus tier

Within the fixed corpus, the papers a system searched were assembled deterministically by a SHA-256-seeded generator keyed by the tuple (query identifier, test type, d), so identical inputs produced the same papers across machines. Test-type composition followed Figure 2 of the main text; the full fill sweep is below (eTable S1.5). The two open-corpus tiers, which re-ask the T4 and T5 questions, replaced the fixed corpus with the live epilepsy PubMed Central pool, restricted to epilepsy by a Medical Subject Headings plus all-fields filter that resolved to 127,076 records (NCBI E-utilities, database pmc, 2026-07-14). The fixed-decompose open port issued one search per sub-question, ranked the whole pool through the NCBI relevance engine, examined the top 20 references, and read up to 3 in full; a one-fact question therefore ranged over up to 20 references and a two-fact question over up to 40. A free-search reference port issued up to 12 search or read actions of 20 results each and reformulated its queries between them.

We denote the distractor fill as d, the resulting number of papers searched (the gold paper(s) plus realized distractors) as N, and the number of evaluated questions as n. N is the gold paper(s) plus distractors: one gold paper for T1 through T4, two for T5. The generator sweeps d rather than N directly, so total N is always d plus the gold count for that test type (d + 1 for T1 through T4, d + 2 for T5). The generator's intended T4 split is about one third near-neighbor to two thirds out-of-domain, filled to the swept target; because the out-of-domain distractor pool is fixed at 980 papers, that ratio cannot hold once d grows large enough to exhaust it, and the generator instead backfills the shortfall with additional near-neighbor papers, up to the size of that pool. eTable S1.5 gives the realized, not intended, composition returned by the generator at every swept d, tabulated directly by consort/realized_haystack_composition.py and checked by unit test; sampling is without replacement throughout, so no paper is searched twice for the same question. T2 and T3 each draw a single distractor type (out-of-domain only and near-neighbor only), and each reaches the end of its pool inside the published sweep. T2 is swept to d = 980, exactly the size of the out-of-domain pool, so it consumes that pool whole without asking for more than it holds (realized N = 981, no shortfall). T3 is swept to d = 1,026 against a near-neighbor pool of 999 non-gold papers, so the request overshoots the pool and is capped: T3's realized maximum is 1,000 papers (999 near-neighbor distractors plus the gold paper), 27 short of the requested 1,027. T5 draws from both pools in an intended half-and-half split but, unlike T4, has no backfill step. At the largest swept d (2,000), the corpus's finite size caps both T4 and T5: the corpus holds 1,980 unique papers in total (1,000 in the gold pool, which also supplies near-neighbor distractors, plus 980 out-of-domain), so no within-corpus N can exceed 1,980 papers regardless of test type. T4’s near-neighbor backfill is itself capped there (999 non-gold papers, all of it), so its realized total (1,980, including its one gold paper) falls 21 short of the requested total of 2,001; T5 has no backfill step and both its near-neighbor and out-of-domain halves are capped at that same d (998 and 980, both pools exhausted), so its realized total (1,980, including its two gold papers) falls 22 short of the requested total of 2,002. These caps sit inside the reported range rather than beyond it. Figure 3 (main text) plots every swept fill, not a reduced set of representative sizes: T1 has one point, the source paper alone; T2 was swept at d = 10, 50, 100, 200, 500 and 980 (realized N = 11 through 981); T3 at d = 10, 50, 100, 200, 500 and 1,026 (realized N = 11 through 1,000); T4 at d = 75, 100, 500, 1,000 and 2,000 (realized N = 76 through 1,980); and T5 at d = 10, 50, 100, 200, 500, 1,000 and 2,000 (realized N = 12 through 1,980). The d = 2,000 rows in eTable S1.5 are therefore reported results and not a structural illustration of the generator’s limit: they are the largest T4 and T5 points in Figure 3, and both were answered against 1,980 papers, the entire corpus, because the corpus is finite and holds no more. The shortfall changes what those points are labeled, not what was measured: each carries its requested fill of 2,000 and was run over 1,980 papers, as T3’s largest point carries 1,026 and was run over 1,000.

**eTable S1.5. Realized T4 and T5 composition of the papers searched: actual generator output at each swept d.** Distractors + gold papers = total N for every row (realized N falls short of the target N only where noted, both at the largest swept d).

| Test type | d (design target) | Gold papers | Near-neighbor realized | Out-of-domain realized | Realized N | Target N (d + gold) |
|---|---:|---:|---:|---:|---:|---:|
| T4 | 75 | 1 | 25 | 50 | 76 | 76 |
| T4 | 100 | 1 | 33 | 67 | 101 | 101 |
| T4 | 500 | 1 | 166 | 334 | 501 | 501 |
| T4 | 1,000 | 1 | 333 | 667 | 1,001 | 1,001 |
| T4 | 2,000 | 1 | 999 | 980 | 1,980 | 2,001 |
| T5 | 75 | 2 | 38 | 37 | 77 | 77 |
| T5 | 100 | 2 | 50 | 50 | 102 | 102 |
| T5 | 500 | 2 | 250 | 250 | 502 | 502 |
| T5 | 1,000 | 2 | 500 | 500 | 1,002 | 1,002 |
| T5 | 2,000 | 2 | 998 | 980 | 1,980 | 2,002 |

T4 meets its target total at every swept d except the largest (2,000): the near-neighbor backfill pool there (999 non-gold papers) is itself smaller than the 1,020 papers a full backfill would require, so T4 also falls 21 papers short there. T5 has no backfill step; at every smaller d in this table it still meets its target because neither half of its intended split exceeds the corresponding pool, but at d = 2,000 both halves are capped (998 non-gold near-neighbor papers, 980 out-of-domain), so T5 falls 22 papers short there. Values are computed with a fixed representative gold paper per query for T1 through T4 and two fixed representative gold papers for T5; realized composition depends only on the pool sizes and the deterministic seed, not on which gold paper(s) are excluded from the near-neighbor pool, so any fixed choice of gold paper(s) is sufficient to tabulate it.

The two refusal tiers invert this construction. For T6 and T7 the gold paper or papers are withheld entirely and the papers searched are drawn only from the 980 non-gold, out-of-domain papers (oncology/endocrinology/cardiology 250, internal medicine 250, surgery/dermatology/infectious disease 249, non-epilepsy neurology 231), swept at d = 1, 10, 50, 200, 500 and 980, where 980 exhausts the pool and is therefore a hard ceiling. The question banks are unchanged: T6 uses all 2,188 reconciled single-paper questions and T7 all 170 two-paper questions, so the refusal tiers ask exactly the questions the retrieval tiers ask, against a corpus that provably cannot answer them.

#### S1.6 Reference harness (IRCoT) and its open port

The reference harness ran a BM25 search over the N papers and retrieved the top 8 passages. It then ran a reason-then-follow-up-query-then-retrieve loop, iterating as long as the model needed more evidence and under a bounded iteration cap, with each round unioning newly retrieved passages into an accumulated set deduplicated by paper identifier and line span. It returned a single grounded answer whose supporting quotes were spliced verbatim from the source body rather than retyped. A distinct-paper ledger enforced the two-distinct-papers requirement of the two-paper tier (T5). The open port applied the same shallow retrieval to the roughly 127,000-record open pool.

The refusal tiers (T6 and T7) used the same IRCoT harness with a single modification: the final-answer instruction permits the system to reply with a fixed refusal marker instead of the usual structured answer. The reason-then-retrieve loop, the retrieval depth (top 8 passages), the iteration cap (3), and the two-fact output contract are byte-identical to the retrieval-tier harness; the permission to decline is the only difference, and it is granted by amending the output-format instruction rather than by appending a clause to it, so the two instructions cannot conflict. The instruction does not state that declining is correct, and does not instruct the system to prefer it.

Responses on the refusal tiers (T6 and T7) were classified into five categories: an explicit refusal, the only credited outcome; a committed fact, incorrect by construction; a hedged answer, which combined a refusal with a substantive clinical claim and received no credit because the two could not be reliably disentangled; a silent non-answer, an empty or unparseable response, which received no credit because it cannot be distinguished from a parsing failure; and an execution error, excluded from the denominators. On T7 a response that declined on one of the two required facts while committing a fact for the other was recorded as a partial refusal and received no credit. The conservative treatment of hedged answers under-counts correct refusals. Two less conservative scoping rules were tried and abandoned because both credited genuine hedges as clean refusals. A response of the form "the papers do not contain any adverse events, confirming safety" was scored as a refusal because the missing-answer and clinical-null-finding senses of "do not contain" could not be separated automatically. DeepSeek-V4-Flash was run over the same refusal grid as the Gemma systems: the Gemma manifest with only the model field swapped, so the work partitions, question set and cell definitions are identical and the arms stay comparable (14,148 cells: 6 values of N × 2,188 single-paper items and 6 × 170 two-paper items). Serving used DeepSeek-V4-Flash-0731 at UD-IQ3_XXS with a 65,536-token context window on 4x NVIDIA L40S per run, with reasoning enabled and a 1,024-token reasoning budget; every run completed with zero failures. Note this is a different build from the panel judge of the same family (DeepSeek-V4-Flash at a 16,384-token context window); the refusal tiers use no language-model judge, so no model contributed to its own score. The matched gold-present control (Table S5b) used the identical serving configuration with the answer-present tier T2 and the deduplicated 1,980-paper corpus, with the reporting paper deliberately kept among the papers searched. Re-deriving outcomes offline from the archived raw answers disagreed with the run-time classification on 56 of 14,148 cells (0.4%): the archived answer text is truncated to 4,000 characters at write time, so a verbose response whose refusal sentinel falls past the cut scores differently offline. The re-derived value is authoritative and the disagreement is flagged per cell in the outcome_mismatch column of the released scorecards.

#### S1.7 Models and serving

The two main Gemma-based systems were Gemma-4B and Gemma-12B at Q8_0 quantization, paired with the IRCoT harness; the models were served locally with llama.cpp (llama-server) through an OpenAI-compatible chat-completions endpoint on NVIDIA L40S graphics processing units. Within-corpus queries used temperature 0.1, top_p 0.95, a fixed random seed of 42, and a 512-token generation limit; the open-corpus agent used temperature 0.2 and a 256-token limit. Model-side reasoning was disabled for Gemma-4B and Gemma-12B through the server reasoning control, because the Gemma chat template otherwise consumed the generation budget on hidden reasoning and returned empty content. The Sonnet-5-based system was Claude Sonnet-5 (identifier claude-sonnet-5) accessed through Claude Code, which performed its own native retrieval over the same corpus rather than the shared IRCoT harness. On the single-paper floor (T1) it read the gold paper in full; on the distractor tiers (T2 to T4) it searched the N papers directly to locate the gold paper among the distractors. Either way it returned one fact with a verbatim supporting quote per question. It therefore differed from the Gemma-based systems in both model and retrieval. The Gemma systems were served from Q8_0 GGUF builds published by Unsloth (gemma-4-E4B-it-Q8_0 and gemma-4-12b-it-Q8_0), each with a 32,768-token context window; Sonnet-5 was accessed through its hosted application programming interface and was not quantized.

#### S1.8 Scoring: three-model panel of judges

The primary metric was a panel of three LLM judges: DeepSeek-V4-Flash, Qwen3.6-35B-A3B, and Llama-4-Scout, served locally. Each judge cast a binary same-fact vote three times, giving nine votes per item, aggregated by per-model majority and then by meta-majority across the three model majorities. The three judges sat outside the roster of evaluated systems, so the panel could not show self-preference;[4] the order of the gold and candidate fields was randomized to control position bias;[3] and model-side reasoning was disabled for the judges. Two defects found during calibration were corrected before the reported runs: the reasoning-enabled judges returned empty structured output that silently scored every cell as incorrect, fixed by disabling judge-side reasoning; and an over-strict rubric that rejected a correct answer for adding detail beyond the gold fact was rewritten to credit the answer when the gold fact’s specific claim was present, marking a cell incorrect only on omission, a different fact, or a contradiction. A deterministic citation-and-quote-overlap score, which credited a candidate only when it cited a gold paper and its supporting quote was recoverable in that paper, was computed for every cell as a byte-reproducible cross-check.

#### S1.9 Statistical analysis

Per-cell accuracy was the proportion of credited questions in a (domain, test type, N) cell. System-level values were arithmetic means over the full N sweep, compared both at matched N and aggregated across N. Because many single-paper questions share a source paper, question-level clustering alone understates single-paper uncertainty; single-paper confidence intervals therefore use a cluster bootstrap by source paper (consort/paper_clustered_ci.py, 5,000 resamples, 835 paper clusters), and single-paper between-system contrasts are paired question-level differences from a question-clustered bootstrap (5,000 resamples drawn by question identifier, so repeated values of N on the same question are not treated as independent; consort/bootstrap_ci.py). For the two-paper set, 95 constituent papers underlie the 170 two-paper questions and a few hub papers place 164 of the 170 pairs in a single connected component (eTable S1.9); a per-question bootstrap would therefore treat the pairs as independent draws when they are not. Two-paper confidence intervals and between-system contrasts instead use a paper-level (graph-aware) bootstrap, which resamples the 95 constituent papers with replacement and carries every pair incident to a resampled paper forward with it, so a pair recurs once for each of its two constituent papers the resample draws (consort/two_hop_graph_ci.py, 5,000 resamples). The comparison was otherwise descriptive and no multiple-comparison correction was applied. Analyses were performed in Python 3.9.6 with NumPy 2.0.2, pandas 2.3.3 and Matplotlib 3.9.4.

eTable S1.9 quantifies that connectivity and checks whether it drives the headline. The 170 two-paper pairs are built from 95 distinct constituent papers, most of which carry more than one pair, and the paper-pair graph has only three connected components, the largest of them holding 164 of the 170 pairs. A component-clustered bootstrap would therefore resample from as few as three quasi-independent units, not 170.Because of this, two-paper confidence intervals use the paper-level (graph-aware) bootstrap described above, which resamples the 95 constituent papers directly rather than the 170 pairs, so the connected-component structure is carried by the resample itself rather than assumed away. As a sensitivity check we excluded every pair touching one of the three papers that each appear in ten or more of the 170 pairs (47 pairs excluded, 123 retained) and recomputed the T5 both-facts headline on the reduced set. Accuracy moved by at most 1.1 points for any system, and every reduced-set estimate fell inside the corresponding full-set confidence interval (lower panel of eTable S1.9), so the reported ordering and magnitude did not depend on the highest-degree papers.

**eTable S1.9. Two-paper paper-pair graph structure and hub-exclusion sensitivity.** Built from the 170 curated two-paper questions and their constituent paper pairs. Connected components are computed over the paper-pair graph (one node per constituent paper, one edge per two-paper question). Both panels' 95% confidence intervals use the paper-level (graph-aware) bootstrap over each panel's own constituent papers; the lower panel repeats the T5 both-facts headline after excluding every pair that touches one of the three papers appearing in ten or more of the 170 pairs.

| Quantity | Value |
|---|---|
| Constituent papers spanning the 170 pairs | 95 |
| Papers appearing in more than one pair | 80 of 95 |

| Quantity | Value |
|---|---|
| Busiest paper’s degree (share of the 170 pairs it supplies a fact for) | 25 (14.7%) |
| Median paper degree | 3 |
| Connected components of the paper-pair graph | 3 |
| Component sizes, largest to smallest (pairs) | 164, 5, 1 |
| Papers excluded in the hub-sensitivity check (degree ≥ 10) | 3 |
| Pairs excluded / retained | 47 / 123 |

| System | T5 both-facts, full set (n = 170) | T5 both-facts, hub-excluded (n = 123) |
|---|---|---|
| Gemma-4B | 5.7% [95% CI 3.6, 8.3] | 6.8% [95% CI 4.1, 10.1] |
| Gemma-12B | 17.1% [95% CI 12.3, 22.4] | 17.1% [95% CI 12.9, 21.4] |
| Sonnet-5 | 37.8% [95% CI 32.5, 43.5] | 37.2% [95% CI 32.3, 42.4] |

Sonnet-5’s single-paper accuracy was measured on the complete reconciled single-paper bank (2,188 questions at each of tiers T1 to T4, 8,752 evaluated cells), not a subsample. Its original July 2026 run covered 2,059 of those items, the 129 remaining items having been the subject of an earlier pilot sweep and therefore omitted from that run’s work list (S1.2); those 129 were regenerated and scored under conditions matched to the 2,059 (same prompt, same native-retrieval harness, and the searched papers rebuilt from the same corpus snapshot and distractor-composition rule), so no single-paper item is now excluded for any system. On generation, Sonnet-5’s answers were verbatim-quote-exact for 96 to 100% of cells and located the correct gold paper among the distractors in 98 to 100% of the T2 to T4 cells. The three-system single-paper head-to-head was computed on the 2,188 questions common to all three systems after reconciliation, at matched N per tier; paired question-clustered differences (Sonnet-5 − Gemma-12B) were +14.5 points (95% CI, 12.8 to 16.3) on T1 and widened to +27.2 points (95% CI, 25.1 to 29.5) on T4 as the two open-weight systems lost accuracy under added distractors while Sonnet-5 did not.

The realized values of N are matched across systems at every tier. T2 and T3 fill to N distractors for every system (realized 101 and 201 papers), and the T4 cells for every system were generated under the same composition rule, which fills to N (realized 501 papers at d = 500; eTable S1.5). The single-paper head-to-head reported above, the T4 contrast included, is therefore a matched-condition comparison, as is every two-paper (T5) comparison.

#### S1.10 Matched two-paper failure decomposition

For the two Gemma-based systems (the Sonnet-5-based system’s decomposition is not reported here), each of the 510 two-paper evaluation cells per system (170 questions across the T5 distractor sweep) was classified by how many of the two gold papers the system retrieved. Both systems retrieved only one of the two gold papers more often than they retrieved both, and retrieved neither in about 1% of cells. Even where both gold papers were retrieved, both facts were credited in a minority of those cells. The joint both-facts accuracy is the retrieve-both rate times this conditional grounding rate, and that product reproduces the reported both-facts value for each system; eTable S1.10 gives every rate.Because the failure to retrieve the second gold paper is larger than the residual failure to ground and combine both facts once retrieved, the dominant two-paper failure mode is second-paper retrieval rather than synthesis alone, with a smaller residual grounding-and-synthesis cost once both papers are found (eTable S1.10). A complementary single-fact isolation baseline for the two-paper constituents was not performed and is not required for the retrieval-versus-grounding decomposition reported here.

**eTable S1.10. Two-paper failure decomposition by paper-retrieval status (Gemma-4B, Gemma-12B).** Each of the 510 evaluated two-paper cells per system (170 questions across the T5 distractor sweep) is classified by how many of the two gold papers were retrieved. The credited-both-given-both row is the conditional grounding-and-synthesis success rate once both papers are found; its product with the retrieve-both rate reproduces the joint both-facts accuracy. The Sonnet-5-based system's decomposition is not reported here.

| Quantity | Gemma-4B | Gemma-12B |
|---|---:|---:|
| T5 cells evaluated | 510 | 510 |
| Retrieved both gold papers | 33.5% | 48.0% |
| Retrieved exactly one gold paper | 65.3% | 51.0% |
| Retrieved neither gold paper | 1.2% | 1.0% |
| Both facts credited, given both papers retrieved | 16.7% | 35.8% |
| Joint both-facts (retrieve-both × credited-given-both) | 5.6% | 17.2% |
| At least one fact credited | 63.9% | 76.2% |
| Two-paper cells credited on neither fact | 36.1% | 23.8% |
| Two-paper cells credited on exactly one fact | 58.3% | 59.1% |
| Two-paper cells credited on both facts | 5.6% | 17.2% |

### S2. Supplementary Tables

**Table S1. Gold-set reconciliation funnel. Counts at each stage of corpus and gold construction, reproduced from Table S1a. The full gold set and the single-paper evaluation bank are distinct artifacts. * Table S1a's footnote applies: this row is the second pass of the manual answerability review in the row above, removing drafts whose gold fact could not answer the question. † Table S1a's footnote applies: two keys carried by the wording-check drop log also appear in the survivor file under a different Unicode normalization and are counted as retained.**

| Stage | Count |
|---|---:|
| Candidate facts extracted | 225,095 |
| Seed-42 stratified sample drawn for | 10,275 |

| Stage | Count |
|---|---|
| review | |
| Not yet reviewed (unprocessed) | 21 |
| Facts approved on first review | 9,728 |
| Facts rejected on first review | 526 |
| Gold facts after two-reviewer reconciliation (256 excluded by the second reviewer) | 9,472 |
| Candidate questions drafted | 9,728 |
| Answerability review, first pass (manual clinician review) | 8,810 |
| Answerability review, second pass (manual clinician review)* | 7,672 |
| No other corpus passage answers them | 5,612 |
| Wording does not give the quote away† | 3,459 |
| Not answerable without reasoning by a small model (fake-reasoning filter) | 2,252 |
| Single-paper evaluation bank, after the gold-fact reconciliation | 2,188 |
| Manually reviewed two-paper questions | 170 |
| Corpus papers (gold + distractor) | 1,980 (1,000 + 980) |

**Table S2. LitBench compared with prior biomedical retrieval and reasoning benchmarks.** For each benchmark, the capability it measures and the property it lacks relative to LitBench.

| Benchmark | What it measures | Property it lacks relative to LitBench |
|---|---|---|
| LitQA2 / PaperQA2 | Identify the correct paper for a free-form question | No gold-quote grounding requirement |
| MIRAGE / MedRAG | Free-form clinical question-answering over PubMed | No gold-quote pool; no graded distractor grid |
| MedCite | Citation accuracy of free-form medical text | Citation only, not retrieval under distraction |
| CRAG | Retrieval-augmented generation across heterogeneous sources | Not biomedical; no source-grounding check |
| BioHopR | Multi-paper biomedical reasoning | No retrieval step; operates on pre-curated facts |
| MedHELM | Broad-coverage medical- | Aggregates other |

| Benchmark | What it measures | Property it lacks relative to LitBench |
|---|---|---|
| | LLM evaluation suite | benchmarks; no source-grounding axis |
| FActScore | Atomic-fact verification of generated text | Measures generation faithfulness, not retrieval |
| ALCE / ASQA | Citation generation in long-form question-answering | Generation operating point, not retrieval under distraction |
| SemioLLM | Seizure-semiology extraction from clinical text | Narrower task; no competing-paper load or multi-paper synthesis |
| PubMedQA / BioASQ | Multiple-choice question-answering over abstracts | Multiple-choice format; no quote grounding |
| SEMQA | Semi-extractive question answering requiring verbatim spans from supplied sources | No retrieval step and no distractor load; not clinical |
| L-CiteEval | Sentence-level citation fidelity of long-context models under swept context length | Padding filtered to minimize overlap with the target, so distractors are non-adversarial; no source-paper credit |
| BrowseComp-Plus | Deep-search agents over a fixed corpus with mined hard negatives | Scores cited document identifiers, not a quoted span; not clinical |
| OpenScholar / ScholarQABench | Retrieval-augmented literature synthesis at open-corpus scale | Citation accuracy judged by entailment, so a paraphrase is credited; no gold required-source set; no distractor-composition control |

**Table S3. Per-configuration, per-N accuracy.** T1 to T5 run from the answering paper alone to two papers with one fact from each. The full per-N grids for every system configuration and the harness comparison of Appendix A are provided as machine-readable per-cell scorecards available from the authors on reasonable request (S5) and are rendered in the figures of S3; they are not reproduced here in full because of their size.

**Table S5. T6/T7 refusal-tier response classification, by model, tier, and N.** For each (model, tier, N) cell: n executed cells and the count and percentage of responses classified as explicit refusal, committed fact, hedged answer, silent non-answer, and execution error (classification defined in the main-text Methods; litbench/core/abstention.py).

| Model | Tier | N | n executed cells | Explicit refusal | Committed fact | Hedged answer | Silent non-answer | Execution error |
|---|---|---|---|---|---|---|---|---|

| | | | | | | | | |
|---|---|---|---|---|---|---|---|---|
| Gemma-12B | T6 | 1 | 2188 | 1932 (88.3%) | 243 (11.1%) | 0 | 13 | 0 |
| Gemma-12B | T6 | 10 | 2188 | 1572 (71.8%) | 616 (28.2%) | 0 | 0 | 0 |
| Gemma-12B | T6 | 50 | 2187 | 1304 (59.6%) | 883 (40.4%) | 0 | 0 | 1 |
| Gemma-12B | T6 | 200 | 2185 | 1079 (49.4%) | 1105 (50.6%) | 0 | 1 | 3 |
| Gemma-12B | T6 | 500 | 2186 | 979 (44.8%) | 1205 (55.1%) | 0 | 2 | 2 |
| Gemma-12B | T6 | 980 | 2181 | 870 (39.9%) | 1311 (60.1%) | 0 | 0 | 7 |
| Gemma-12B | T7 | 1 | 170 | 158 (92.9%) | 12 (7.1%) | 0 | 0 | 0 |
| Gemma-12B | T7 | 10 | 170 | 129 (75.9%) | 41 (24.1%) | 0 | 0 | 0 |
| Gemma-12B | T7 | 50 | 169 | 111 (65.7%) | 58 (34.3%) | 0 | 0 | 1 |
| Gemma-12B | T7 | 200 | 170 | 94 (55.3%) | 76 (44.7%) | 0 | 0 | 0 |
| Gemma-12B | T7 | 500 | 170 | 76 (44.7%) | 94 (55.3%) | 0 | 0 | 0 |
| Gemma-12B | T7 | 980 | 170 | 68 (40.0%) | 102 (60.0%) | 0 | 0 | 0 |
| Gemma-4B | T6 | 1 | 2188 | 9 (0.4%) | 2172 (99.3%) | 0 | 7 | 0 |
| Gemma-4B | T6 | 10 | 2186 | 0 (0.0%) | 2183 (99.9%) | 0 | 3 | 2 |
| Gemma-4B | T6 | 50 | 2186 | 0 (0.0%) | 2182 (99.8% | 0 | 4 | 2 |

| | | | | | | | | |
|---|---|---|---|---|---|---|---|---|
| | | | | | ) | | | |
| Gemma-4B | T6 | 200 | 2186 | 0 (0.0%) | 2184 (99.9%) | 0 | 2 | 2 |
| Gemma-4B | T6 | 500 | 2184 | 0 (0.0%) | 2181 (99.9%) | 0 | 3 | 4 |
| Gemma-4B | T6 | 980 | 2187 | 0 (0.0%) | 2184 (99.9%) | 0 | 3 | 1 |
| Gemma-4B | T7 | 1 | 170 | 0 (0.0%) | 170 (100.0%) | 0 | 0 | 0 |
| Gemma-4B | T7 | 10 | 170 | 0 (0.0%) | 169 (99.4%) | 0 | 1 | 0 |
| Gemma-4B | T7 | 50 | 170 | 0 (0.0%) | 170 (100.0%) | 0 | 0 | 0 |
| Gemma-4B | T7 | 200 | 170 | 0 (0.0%) | 170 (100.0%) | 0 | 0 | 0 |
| Gemma-4B | T7 | 500 | 170 | 0 (0.0%) | 170 (100.0%) | 0 | 0 | 0 |
| Gemma-4B | T7 | 980 | 170 | 0 (0.0%) | 170 (100.0%) | 0 | 0 | 0 |
| DeepSeek-V4-Flash | T6 | 1 | 2188 | 2185 (99.9%) | 0 (0.0%) | 3 | 0 | 0 |
| DeepSeek-V4-Flash | T6 | 10 | 2188 | 2075 (94.8%) | 88 (4.0%) | 20 | 5 | 0 |
| DeepSeek-V4-Flash | T6 | 50 | 2188 | 1952 (89.2%) | 185 (8.5%) | 37 | 14 | 0 |
| DeepSeek-V4-Flash | T6 | 200 | 2188 | 1849 (84.5%) | 286 (13.1%) | 33 | 20 | 0 |
| DeepSeek-V4-Flash | T6 | 500 | 2188 | 1739 (79.5%) | 361 (16.5%) | 45 | 43 | 0 |
| DeepSeek-V4-Flash | T6 | 980 | 2188 | 1704 (77.9%) | 407 (18.6% | 47 | 30 | 0 |

| | | | | | | | | |
|---|---|---|---|---|---|---|---|---|
| | | | | | ) | | | |
| DeepSeek-V4-Flash | T7 | 1 | 170 | 170 (100.0%) | 0 (0.0%) | 0 | 0 | 0 |
| DeepSeek-V4-Flash | T7 | 10 | 170 | 169 (99.4%) | 1 (0.6%) | 0 | 0 | 0 |
| DeepSeek-V4-Flash | T7 | 50 | 170 | 166 (97.6%) | 2 (1.2%) | 1 | 1 | 0 |
| DeepSeek-V4-Flash | T7 | 200 | 170 | 164 (96.5%) | 4 (2.4%) | 1 | 1 | 0 |
| DeepSeek-V4-Flash | T7 | 500 | 170 | 155 (91.2%) | 8 (4.7%) | 4 | 3 | 0 |
| DeepSeek-V4-Flash | T7 | 980 | 170 | 158 (92.9%) | 6 (3.5%) | 4 | 2 | 0 |

**Table S5b. Gold-present control arm for DeepSeek-V4-Flash (T2). The same abstention-permitting instruction and the same harness as Table S5, but the paper reporting the answer is among the papers searched, so an explicit refusal is a false refusal.**This is the floor against which the T6/T7 rates in Table S5 must be read: a system that declines to most questions scores highly wherever declining is correct. 300 cells, using the same harness implementation as the abstention arm, configured to run against T2 instead of the refusal tiers.

| N (papers searched) | n executed cells | False refusal | Committed fact | Hedged answer |
|---|---|---|---|---|
| 10 | 50 | 47 (94.0%) | 1 | 1 |
| 50 | 50 | 45 (90.0%) | 4 | 1 |
| 100 | 50 | 39 (78.0%) | 8 | 3 |
| 200 | 50 | 39 (78.0%) | 8 | 3 |
| 500 | 50 | 35 (70.0%) | 10 | 5 |
| 980 | 50 | 36 (72.0%) | 13 | 1 |
| All | 300 | 241 (80.3%) | 44 | 14 |

**Table S6. Worked example from the two-paper question bank (query M30073).** The example contrasts complete and incomplete evidence retrieval across tested values of N. Aggregate results are in main-text Table 2 and Figure 3E.

| | |
|---|---|
| Clinical question | A language-dominant temporal tumor carries oncologic progression risk but resection threatens speech. Which exact operative strategy should merge tumor control with language preservation? |
| Reference fact A (Delev et al., Sci Rep 2020) | Patients with LEATs showing astrocytic differentiation, BRAF mutation, or MAPK/FGFR/EGFR pathway activation are at |

| | |
|---|---|
| | higher risk of tumor recurrence and malignant progression; treatment should prioritize gross total resection from an oncological standpoint. |
| Reference fact B (Serrano-Castro et al., Front Neurol 2020) | Direct cortical stimulation mapping and language pre-habilitation before epilepsy surgery can rescue patients otherwise excluded from resection, especially younger patients with long-standing lesions. |
| Sonnet-5 (cloud-hosted commercial model) | Credited on both facts at N=10, 50, and 200, with near-verbatim quotations matching both reference passages; both-fact credit was not consistent across all tested values of N (main-text Figure 3E). |
| Gemma-4B (open-weight model that can run entirely on a hospital's own hardware) | Not credited on both facts together at any tested value of N (N=10, 50, or 200). |

Refusal counts and percentages are of executed cells (scored), which excludes execution errors. Hedged responses (a refusal combined with a substantive clinical claim) earn no credit and were zero in all 28,296 cells, so the conservative classification rule did not suppress any refusal. Silent non-answers (empty or unparseable output) earn no credit and never count as abstention. Partial refusals (one of the two required facts declined and a fact committed for the other) numbered 2, both in the Gemma-12B two-paper refusal tier (T7). Sonnet-5 was evaluated on a sample of these tiers (Appendix B).

### S3. Supplementary Figures

The refusal-tier data appear in the main text as Figure 3, panels F and G. This section collects the per-domain grids (Figure S1), the combined two-paper surface across every distractor fill (Figure S2), the single-paper versus two-paper comparison (Figure S3), the harness screen (Figure A1) and the harness itself (Figure A2); the underlying per-cell values are those summarized in Table S3.

**Figure A1. Gemma-4B two-paper accuracy of each candidate retrieval harness across the full sweep of N**, the harness screen underlying Appendix A

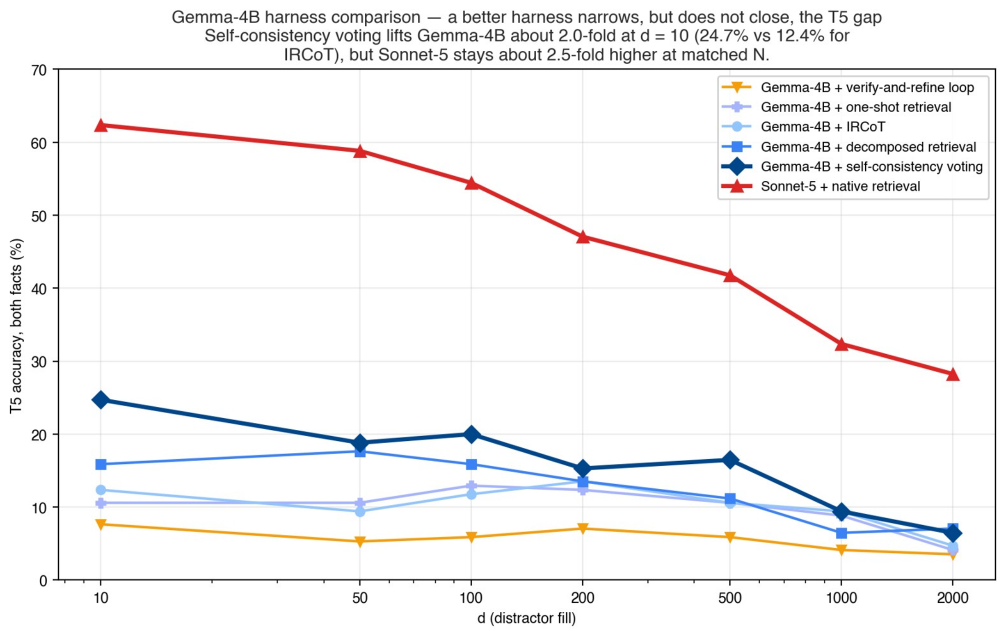


**Figure A2. The IRCoT retrieval harness: one bounded reason-then-retrieve loop**, shared by both Gemma configurations and applied to the single-paper tiers, from the answering paper alone (T1) to hidden among both kinds of competing paper (T4), and the two-paper tier, two papers with one fact from each (T5)

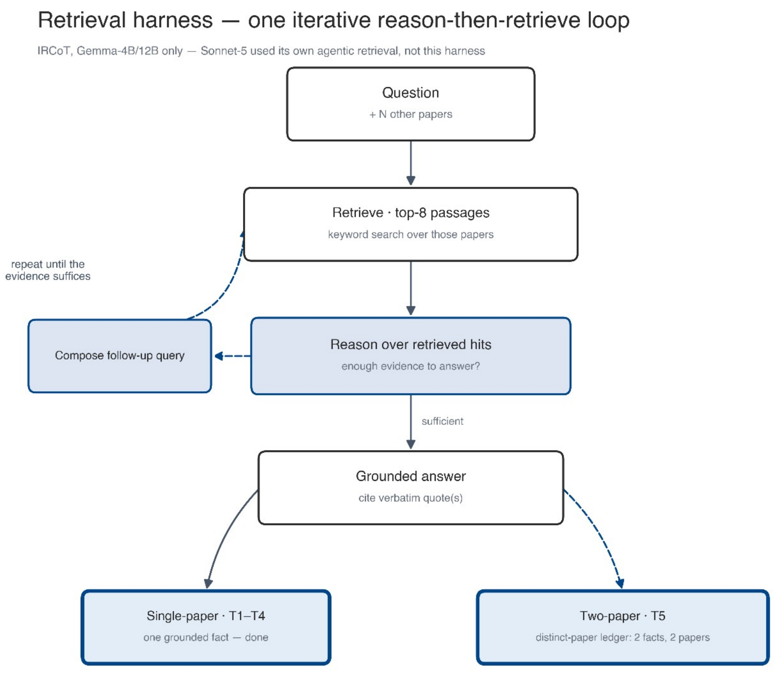


**Figure S1. Full within-corpus performance grid for the IRCoT systems and Sonnet-5's native retrieval, shown in two panels.** The three model bands are split across two panels (upper: Gemma-4B and Gemma-12B; lower: Sonnet-5) purely for legibility; the grid, data and color scale are identical and nothing is omitted between them. Panel accuracy (%) of the three-model judge panel (DeepSeek-V4-Flash, Qwen3.6-35B-A3B, Llama-4-Scout), shown for every cell of the benchmark. The three colored bands are the three main systems (Gemma-4B and Gemma-12B with the IRCoT harness, Sonnet-5 with its native retrieval); within each band, columns step through the full distractor-fill sweep of each difficulty tier (test type × fill, T1 through the two-paper T5 and the open-corpus ∞ tail); rows are the 14 fact domains (seven article sections crossed with the two fact types, stated outright and requiring interpretation) plus an aggregate row. The color scale runs continuously from red (lower accuracy) through amber to green (higher accuracy); white = a tier that is structurally absent for that domain (the methods, table, and figure interpretation-requiring domains carry no two-paper questions by construction). This is the full, fully populated three-band grid, and the per-domain exhibit the main text points to; it adds the distractor-fill sweep and the open-corpus tail that main-text Figure 1 does not carry. Main-text Figure 1 shows the same domains across all seven conditions, including the two refusal tiers and the fourth system. Per-cell values are set for on-screen reading and are best inspected by zooming the vector PDF

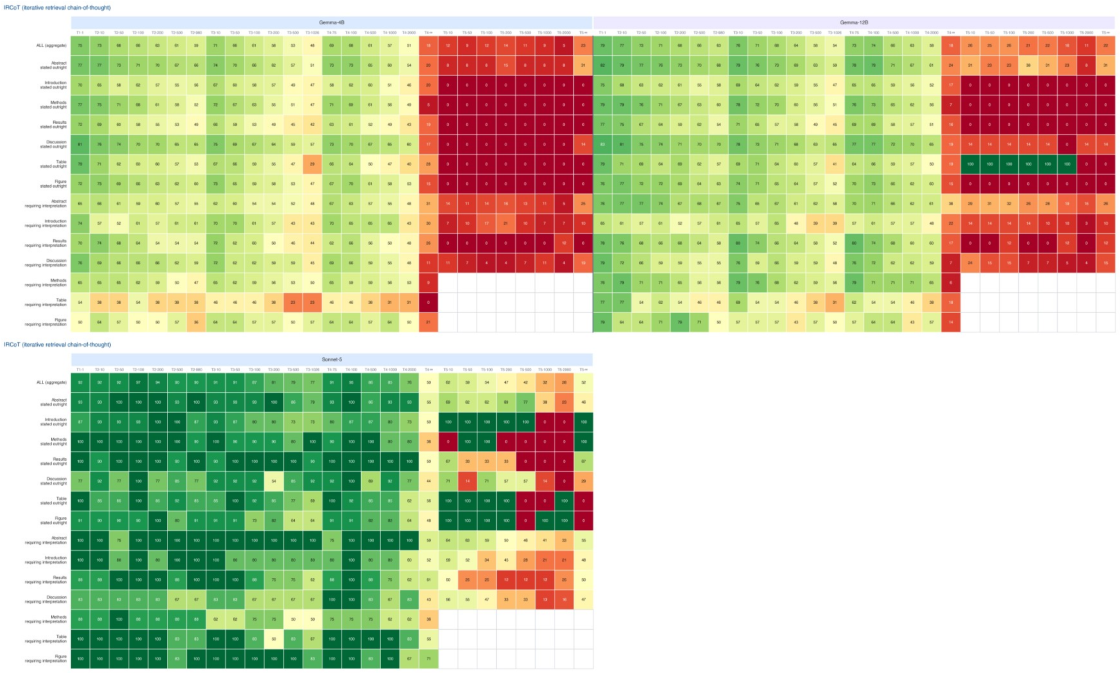


**Figure S2. Combined full two-paper (T5) surface**, every system as a row and each distractor fill d as a column; three-model judge panel (DeepSeek-V4-Flash, Qwen3.6-35B-A3B, Llama-4-Scout) both-facts accuracy on the 170 manually reviewed two-paper queries.

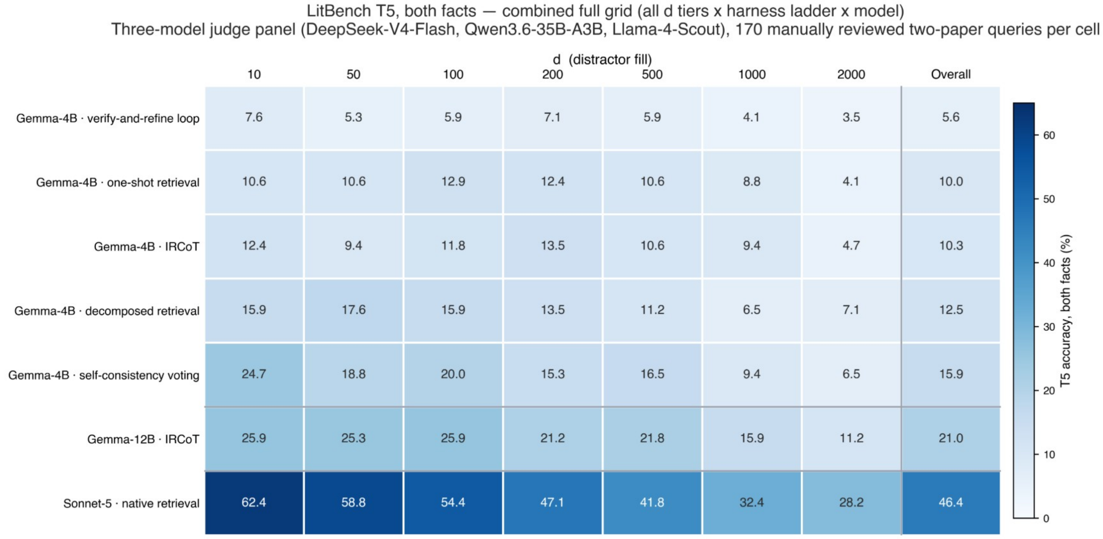


**Figure S3. Single-item-set versus two-paper-item-set accuracy (descriptive).** For each of the three main systems (Gemma-4B + IRCoT, Gemma-12B + IRCoT, Sonnet-5 + native retrieval, each in its own color), the solid bar is the single-paper mean accuracy across tiers T1 to T4 (matched n = 2,188 questions) and the hatched bar is two-paper accuracy on T5 (both gold facts required, pooled across the full sweep of N); both are three-model judge panel accuracies. Error bars on the single-paper bars are paper-clustered bootstrap 95% confidence intervals (835 gold papers); error bars on the two-paper bars are paper-level (graph-aware) bootstrap 95% confidence intervals (95 constituent papers underlying the 170 pairs). Δ (percentage points) is annotated above each pair. The single-paper bar draws on one gold paper's questions and the two-paper bar on a two-paper synthesis question, so this is a descriptive comparison of different item sets rather than a paired before/after measurement on the same items.

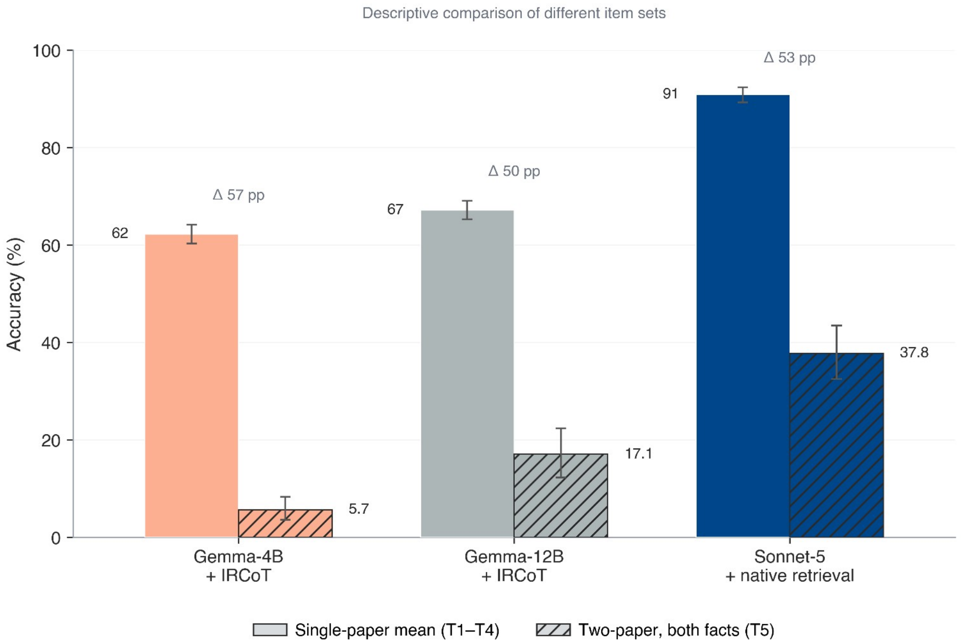


### S4. Judge-Validation Protocol

#### S4.1 Published panel-of-judges evidence

The panel design follows published evidence that a panel of diverse LLM judges aligns with human labels at least as well as a single strong judge,[1] at the binary multi-paper operating point that matches LitBench. Published guidance also recommends validating an LLM judge against human labels before deployment,[2] which motivates the benchmark-specific protocol below.[5]

**Table S4. Published LLM-judge agreement with human labels.** Cohen κ of a panel of LLMs and of a single GPT-4 judge against human labels on question-answering benchmarks, and the human-agreement anchor from MT-Bench.

| Source | Benchmark | Panel-of-LLMs κ | Single-judge κ |
|---|---|---:|---:|
| Verga et al. 2024 | HotpotQA (multi-paper) | 0.867 | 0.830 |
| Verga et al. 2024 | Bamboogle (multi-paper) | 0.896 | not reported |
| Verga et al. 2024 | TriviaQA | 0.906 | 0.841 |
| Verga et al. 2024 | Natural Questions | 0.763 | 0.627 |
| Zheng et al. 2023 (MT-Bench) | Human-preference | over 80% (human-human comparable) | not applicable |

| Source | Benchmark agreement | Panel-of-LLMs κ | Single-judge κ |
|---|---|---|---|

#### S4.2 Single-reviewer validation results

The first author (A.G.), a physician, labeled a validation sample of the judge panel's cells (n = 154 cells: 26 T1 and 32 each of T2 to T5, running from the answering paper alone to two papers with one fact from each), recording for each cell whether the returned candidate fact conveyed the same clinical claim as the gold fact, without consulting the panel's verdict. Each label was joined to the judge panel's credit for the same cell, with the two-paper cells scored per-paper (panel majority on each gold fact, requiring both) exactly as in the main results. The judge panel agreed with the clinician on 89.0% of the 154 cells: Cohen κ 0.774 (95% CI 0.67 to 0.87), Gwet AC1 0.784, sensitivity 0.93, and specificity 0.84 (confusion: 80 true-positive, 6 false-negative, 57 true-negative, 11 false-positive). Per-tier agreement follows (Table S4b). Disagreements were slightly more often over-credits than misses (11 versus 6 of 154), so the panel erred marginally toward crediting rather than under-crediting.

**Table S4b. Panel-clinician agreement by test type (n = 154 cells, single reviewer: the first author).** Cohen κ and Gwet AC1 with per-tier n; sensitivity and specificity take the clinician label as reference. Two-paper (T5) cells were scored per-paper (panel majority on each gold fact, requiring both), matching the main-results scorer.

| Test type | n | Raw agreement | Cohen κ | Gwet AC1 | Sensitivity | Specificity |
|---|---|---|---|---|---|---|
| T1 | 26 | 92.3% | 0.80 | 0.87 | 0.95 | 0.86 |
| T2 | 32 | 90.6% | 0.80 | 0.82 | 0.95 | 0.85 |
| T3 | 32 | 87.5% | 0.72 | 0.77 | 0.95 | 0.75 |
| T4 | 32 | 84.4% | 0.69 | 0.69 | 0.88 | 0.81 |
| T5 | 32 | 90.6% | 0.80 | 0.82 | 0.92 | 0.90 |
| Overall | 154 | 89.0% | 0.77 | 0.78 | 0.93 | 0.84 |

The lowest per-tier estimate is T4 (κ 0.69, n = 32); each per-tier estimate is small enough that a few reclassified cells shift κ by roughly 0.1. Across the sample the panel's disagreements were predominantly over-credits (11 of 17), consistent with the marginally lenient direction noted above.

#### S4.3 Deterministic cross-check

The reported accuracies rely on the judge panel anchored to the published evidence in Table S4 and the single-reviewer validation above (S4.2), and on the deterministic citation-and-quote-overlap cross-check computed for every cell (S1.8). Table S4c reports this cross-check by system and condition.

**Table S4c. Composite span-checked endpoint, by system and condition.** The secondary endpoint reported alongside the panel credit of main-text Table 2. Credit requires the correct source paper AND a supporting span reproduced verbatim from it AND panel confirmation that the returned fact conveys the same clinical claim. It is deterministic given the run records and uses no language model, so it is byte-reproducible; it scores lower than panel credit by construction, because a system that paraphrases a real supporting sentence loses the span test.

| System | T1 gold paper alone | T2 + unrelated | T3 + similar | T4 + both | T5 both facts |
|---|---|---|---|---|---|
| Gemma-4B + IRCoT | 63.9 (61.6–66.2) | 57.4 (55.1–59.7) | 49.2 (47.0–51.5) | 52.3 (49.9–54.6) | 5.1 |
| Gemma-12B + IRCoT | 67.4 (65.2–69.5) | 61.1 (58.9–63.2) | 55.0 (52.7–57.2) | 59.2 (56.9–61.4) | 15.3 |
| Sonnet-5 + native retrieval (Claude Code) | 88.3 (86.5–89.9) | 86.3 (84.5–88.0) | 86.0 (84.2–87.8) | 85.8 (84.0–87.6) | 26.0 |

### S5. Code and Data Availability

The list of the 1,980 corpus articles is published: title, journal, year, digital object identifier and a resolvable link for each, with the 1,000 epilepsy papers that carry the answers marked apart from the 980 non-epilepsy distractors that carry none. It is publicly available on GitHub (https://github.com/GoldenholzLab/LitBench), together with the analysis and figure-generation code that produces the reported confidence intervals and between-system contrasts from bundled aggregate result files. The benchmark construction pipeline, the retrieval harness and the abstention parser are not in that repository. The benchmark itself is not published. To keep LitBench an uncontaminated test set and out of future model training, the question bank, the gold facts, the per-cell scorecards for every system configuration and test condition, and the candidate and judge prompts are withheld. That split is what makes publishing the article list safe: the list says which papers a system had to search, not which facts were asked about or what the correct answers are. The withheld materials are available from the authors on reasonable request. The deterministic citation-and-quote-overlap score can be recomputed from the provided run records without any external service. The judge and candidate model weights are the open-weight and proprietary models named in the main-text Methods.

### References


1. Verga P, Hofstatter S, Althammer S, et al. Replacing judges with juries: evaluating LLM generations with a panel of diverse models. arXiv preprint arXiv:2404.18796. 2024.
2. Zheng L, Chiang WL, Sheng Y, et al. Judging LLM-as-a-judge with MT-Bench and Chatbot Arena. In: *Advances in Neural Information Processing Systems 36: Datasets and Benchmarks Track*. 2023.
3. Wang P, Li L, Chen L, et al. Large language models are not fair evaluators. In: *Proceedings of the 62nd Annual Meeting of the Association for Computational*

*Linguistics (Volume 1: Long Papers)*. 2024:9440-9450. doi:10.18653/v1/2024.acl-long.511

4. Panickssery A, Bowman SR, Feng S. LLM evaluators recognize and favor their own generations. In: *Advances in Neural Information Processing Systems 37*. 2024.
5. Bavaresco A, Bernardi R, Bertolazzi L, et al. LLMs instead of human judges? A large scale empirical study across 20 NLP evaluation tasks. In: *Proceedings of the 63rd Annual Meeting of the Association for Computational Linguistics (Volume 2: Short Papers)*. 2025:238-255. doi:10.18653/v1/2025.acl-short.20
